# Genetic Associations with Placental Proteins in Maternal Serum Identify Biomarkers for Hypertension in Pregnancy

**DOI:** 10.1101/2023.05.25.23290460

**Authors:** Qi Yan, Nathan R. Blue, Buu Truong, Yu Zhang, Rafael F. Guerrero, Nianjun Liu, Michael C. Honigberg, Samuel Parry, Rebecca B. McNeil, Hyagriv N. Simhan, Judith Chung, Brian M. Mercer, William A. Grobman, Robert Silver, Philip Greenland, George R. Saade, Uma M. Reddy, Ronald J. Wapner, David M. Haas

## Abstract

**Background:** Preeclampsia is a complex syndrome that accounts for considerable maternal and perinatal morbidity and mortality. Despite its prevalence, no effective disease-modifying therapies are available. Maternal serum placenta-derived proteins have been in longstanding use as markers of risk for aneuploidy and placental dysfunction, but whether they have a causal contribution to preeclampsia is unknown.

**Objective:** We aimed to investigate the genetic regulation of serum placental proteins in early pregnancy and their potential causal links with preeclampsia and gestational hypertension.

**Study design:** This study used a nested case-control design with nulliparous women enrolled in the nuMoM2b study from eight clinical sites across the United States between 2010 and 2013. The first- and second-trimester serum samples were collected, and nine proteins were measured, including vascular endothelial growth factor (VEGF), placental growth factor, endoglin, soluble fms-like tyrosine kinase-1 (sFlt-1), a disintegrin and metalloproteinase domain-containing protein 12 (ADAM-12), pregnancy-associated plasma protein A, free beta-human chorionic gonadotropin, inhibin A, and alpha-fetoprotein. This study used genome-wide association studies to discern genetic influences on these protein levels, treating proteins as outcomes. Furthermore, Mendelian randomization was used to evaluate the causal effects of these proteins on preeclampsia and gestational hypertension, and their further causal relationship with long-term hypertension, treating proteins as exposures.

**Results:** A total of 2,352 participants were analyzed. We discovered significant associations between the pregnancy zone protein locus and concentrations of ADAM-12 (rs6487735, *P=*3.03×10^-22^), as well as between the vascular endothelial growth factor A locus and concentrations of both VEGF (rs6921438, *P=*7.94×10^-30^) and sFlt-1 (rs4349809, *P=*2.89×10^-12^). Our Mendelian randomization analyses suggested a potential causal association between first-trimester ADAM-12 levels and gestational hypertension (odds ratio=0.78, *P=*8.6×10^-4^). We also found evidence for a potential causal effect of preeclampsia (odds ratio=1.75, *P*=8.3×10^-3^) and gestational hypertension (odds ratio=1.84, *P*=4.7×10^-3^) during the index pregnancy on the onset of hypertension 2-7 years later. The additional mediation analysis indicated that the impact of ADAM-12 on postpartum hypertension could be explained in part by its indirect effect through gestational hypertension (mediated effect=-0.15, *P=*0.03).

**Conclusions:** Our study discovered significant genetic associations with placental proteins ADAM-12, VEGF, and sFlt-1, offering insights into their regulation during pregnancy. Mendelian randomization analyses demonstrated evidence of potential causal relationships between the serum levels of placental proteins, particularly ADAM-12, and gestational hypertension, potentially informing future prevention and treatment investigations.

## Introduction

Preeclampsia (PE) is a complex syndrome of widespread maternal endothelial activation and intravascular inflammation with a range of contributing factors arising from a final common pathway of dysfunction at the maternal-placental interface, with progressive clinical deterioration unless placental delivery is achieved.^1^ Despite considerable etiologic complexity, the placenta appears to be central to the development of PE, either as the primary pathological source or as a secondary affected organ in the setting of other insults.^2^ Because the perturbations leading to PE occur in early pregnancy and are followed by a long pre-clinical phase that precedes clinical signs, prior efforts have focused on early, non-invasive detection of placental dysfunction as a means for clinical prediction.^3–6^ We previously analyzed clinical and biospecimen data from the Nulliparous Pregnancy Outcomes Study: Monitoring Mothers-to-Be (nuMoM2b) cohort, in which we found that the maternal serum levels of nine placenta-derived proteins collected during the first and second trimesters (n=2,352) were associated with PE.^7^ Other studies also have found that circulating levels of placental proteins, particularly angiogenic factors produced by the placenta, are associated with PE.^3, 4, 8–10^

Beyond the short-term implications of PE, there is increasing recognition of its long-term maternal associations with later-life hypertension, cardiovascular (CV) disease, and renal disease.^11–15^ In this setting, the incidence of PE or gestational hypertension (gHTN) serves as an indicator of future CV risk, but it remains unclear whether the PE/gHTN is causal or is merely an indicator of underlying genetic predisposition to CV disease.

Addressing these knowledge gaps in PE care may be possible through interrogation of the maternal genome using an approach called Mendelian randomization.^16^ This approach, which uses genetic variants randomly assigned at conception as proxies for risk factors, has become increasingly popular in clinical research, mimicking a randomized clinical trial.^17–19^ Such an approach has the potential to identify a biomarker as a therapeutic target or clarify that PE may cause rather than merely predict future CV risk.

To fill the knowledge gap, we first set out to clarify the genetic influences on early pregnancy placental protein levels, which had previously not been done during pregnancy.^20–26^ Thus, we performed genome-wide association studies (GWAS) on nine placental protein levels measured in maternal serum during the first (visit1) and second (visit2) trimesters, as well as the change between the two time points (visit2-1), among nuMoM2b participants. Identifying protein-genetics associations can pinpoint disease linked proteins influenced by genetic variations, thereby facilitating novel therapeutic target identification.^27, 28^ We then carried out Mendelian randomization, based on the GWAS findings for these nine proteins and the most recent large GWAS on PE,^29^ to investigate whether maternal serum placental protein levels in early pregnancy are potential causal factors for PE and gHTN. Finally, we used Mendelian randomization to examine the potential causal relationship between PE/gHTN and long-term maternal HTN.

## Materials and Methods

The nuMoM2b study was designed as a prospective cohort study aimed at investigating the underlying causes and pathophysiological pathways associated with adverse pregnancy outcomes (APOs) in nulliparous pregnant women. The objective of this study is to identify the associations between placental proteins and genetic variations, and to further explore their roles in causing PE, gHTN, and long-term HTN. A subsample of the nuMoM2b cohort was used to perform the GWAS of placental proteins. Leveraging external large-scale GWAS data on PE/gHTN,^29^ combined with nuMoM2b data, Mendelian randomization was employed to investigate the causal relationships. Details of the data and analysis are described in the sections that follow.

## Cohorts

The nuMoM2b study enrolled 10,038 women from eight geographically disparate clinical sites in the United States and was designed to recruit a large, racially, and geographically diverse cohort of nulliparous pregnant women (Supplemental Table 1).^30^ The study participants were longitudinally followed and underwent four study visits, from the first trimester to after birth. Throughout pregnancy, various data were obtained, including detailed interviews, questionnaires, research ultrasounds, maternal biometric measurements, and biospecimens (Supplemental Table 2). Peripheral maternal blood samples were collected at three study visits during pregnancy: visit1 (6-13 weeks), visit2 (16-21 weeks), and visit3 (22-29 weeks). The methods of the nuMoM2b study have been described in detail elsewhere,^30^ and the study was approved by the Institutional Review Boards at all participating centers.^30–32^ Genome-wide genotyping of 9,757 women with sufficient material was performed with whole blood samples collected at visit1, using the Infinium Multi-Ethnic Global D2 BeadChip (Illumina, Miami, USA). The nuMoM2b Heart Health Study (nuMoM2b-HHS) was carried out as a sequel study to gain a better understanding of the influence of pregnancy outcomes on subsequent health,^32^ and 4,484 women completed the laboratory assessments at the 2-7 year postpartum in-person visit (Supplemental Table 2).

Maternal serum samples were collected at visit1 and visit2 to measure the levels of nine placental proteins, including vascular endothelial growth factor (VEGF) (pg/mL), placental growth factor (PlGF) (pg/mL), endoglin (ENG) (ng/mL), soluble fms-like tyrosine kinase-1 (sFlt-1) (pg/mL), a disintegrin and metalloproteinase domain-containing protein 12 (ADAM-12) (ng/mL), pregnancy-associated plasma protein A (PAPP-A) (mU/mL), free beta-human chorionic gonadotropin (fβHCG) (ng/mL), inhibin A (INHA) (pg/mL), and alpha fetal protein (AFP) (IU/mL). They fall into three categories that reflect different aspects of placental function: (1) angiogenesis (VEGF, PlGF, ENG, and sFlt-1);^4, 8, 33–38^ (2) placental implantation and development (ADAM-12 and PAPP-A);^39, 40^ and (3) biomarkers for fetal chromosomal abnormalities (fβHCG, INHA, and AFP).^41, 42^ While AFP does not originate from the placenta, its increased levels in maternal serum during the second trimester are associated with APOs, likely linked to excessive placental permeability.^43, 44^ The analysis was conducted in a subsample of the nuMoM2b cohort with available genotypes (n=2,352), which included women who experienced any of the following APOs (n=1,463): delivery prior to 37 weeks’ gestation, PE or eclampsia, birth weight for gestational age <5th percentile, or stillbirth. It also included 889 controls who delivered at term without complications. The protein levels were log-transformed for subsequent analyses.

## Analytical workflow for quality control and genome-wide association study with multi-ethnic data

In multi-ethnic GWAS, it is common to split the total sample into separate populations, perform genotype data quality control (QC), imputation, and GWAS in each population separately, followed by a meta-analysis. However, in our study, the sample size for GWAS of protein levels is relatively small, at around two-thousand individuals. Further splitting the data into separate populations would make GWAS unreasonable for certain populations with limited sample sizes. The principal component analysis (PCA) revealed a continuous population structure in the nuMoM2b cohort (Supplemental Figure 1). Consequently, we have developed an analytical workflow for QC and GWAS with multi-ethnic data that maximizes the sample size while minimizing bias due to population stratification (Supplemental Figure 2). Genotype imputation was performed using the TOPMed Imputation Server,^45^ retaining genotyped and imputed SNPs with imputation quality r^2^>0.3. Our refined nuMoM2b cohort included 9,742 women. In the protein GWAS, 2,263, 2,134, and 2,045 women were analyzed for visit1, visit2, and visit2-1 analyses, respectively. Given our sample size, we concentrated on SNPs with MAF>0.05. To control for population stratification, we used the GENESIS R/Bioconductor package^46, 47^ to fit linear mixed models that integrated a random effect to control for genetic relatedness. The covariates included age and age-squared at visit1, first 10 principal components (PCs) calculated by genotypic data, self-reported race, clinical sites, and status for any APOs. Additionally, we conducted supplementary genome-wide interaction studies (GWIS) to remove SNPs with different effects across ancestries. We also ensured no impact from collider bias in our analyses. A genome-wide significance threshold was set at *P<*5.6×10^-9^ (Bonferroni-adjusted for nine proteins: 5×10^-8^/9=5.6×10^-9^). This analytical workflow is detailed in the supplemental text.

## Causal inference

Mendelian randomization uses SNPs as instrumental variables (IVs) to estimate the causal effect of a risk factor on the outcome of interest, while removing unmeasured confounding bias. The design of our Mendelian randomization study for causal inference is depicted in Figure 1A, which includes three analyses: 1) proteins → PE/gHTN, 2) PE/gHTN → long-term postpartum HTN, and 3) proteins → PE/gHTN → long-term HTN. Several large-scale GWAS have been conducted on PE.^29, 48, 49^ Our study leverages the most recent large GWAS,^29^ analyzing PE with 17,150 cases and 451,241 controls, as well as gestational hypertension (gHTN) with 8,961 cases and 184,925 controls. As with any Mendelian randomization analysis, some assumptions, such as horizontal pleiotropy, are untestable. To address this, employing multiple approaches with different assumptions and achieving consistent results can enhance confidence in causal effect estimates. In this study, we considered three commonly used approaches to ensure result robustness. Our primary method was the Mendelian randomization-robust adjusted profile scoring (MR-RAPS)^50^, given its capability to adjust for weak instrument bias which is particularly relevant in our study with a limited number of strong IV SNPs. We also used random-effect inverse variance weighting (IVW),^51^ and Mendelian randomization pleiotropy residual sum and outlier (MR-PRESSO)^52^ as sensitivity analyses. We set the threshold at *P<*5.6×10^-3^, Bonferroni-adjusted for nine proteins. The two Mendelian randomization analyses, proteins → PE/gHTN and PE/gHTN → long-term postpartum HTN, were used for mediation analysis using the product of coefficients method to estimate the mediated effect for the pathway from proteins → PE/gHTN → long-term postpartum HTN.^53^ Details of Mendelian randomization analyses can be found in the supplemental text.

**Figure 1.**
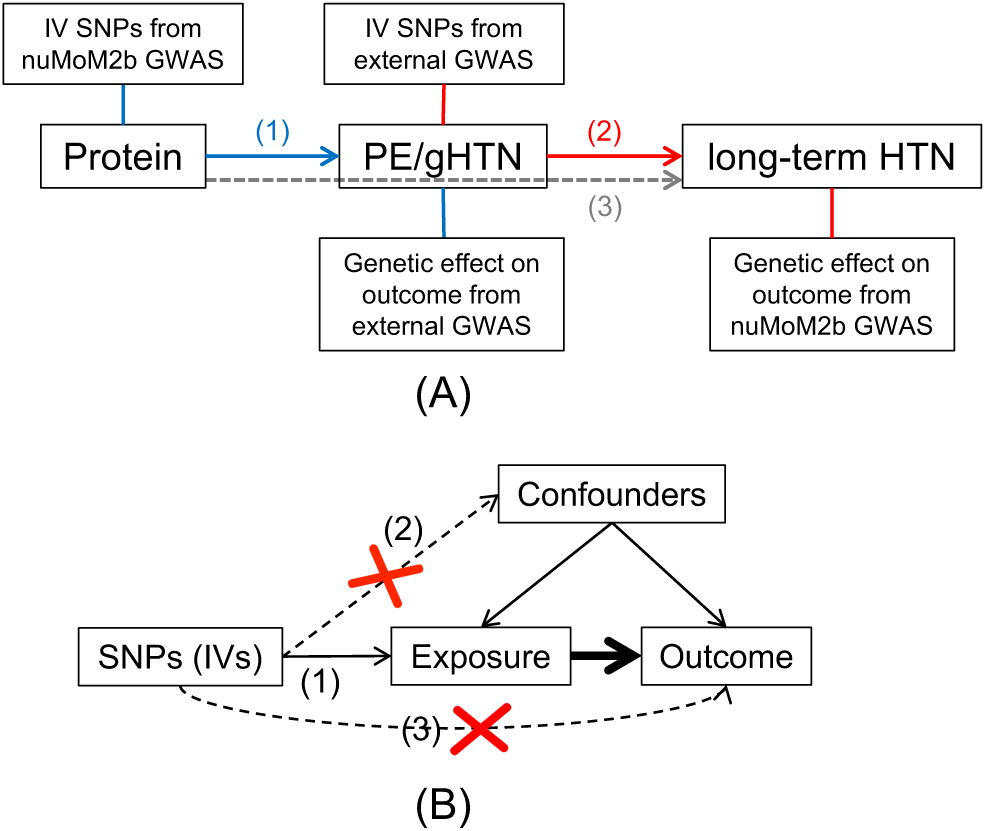
Mendelian randomization for causal inference. (A) Flowchart for two-sample Mendelian randomization. (1) Causal relationship between placental protein levels and PE/gHTN; (2) Causal relationship between PE/gHTN and long-term postpartum HTN; and (3) Causal mediation effects of placental protein levels on long-term postpartum HTN via PE/gHTN. (B) Illustrative diagram of Mendelian randomization. Its validity relies on 3 assumptions: IVs need to be (1) associated with the exposure, (2) not associated with any confounder of the exposure-outcome association, and (3) independent of the outcome conditional on the exposure and confounders (the horizontal pleiotropy assumption).

## Results

### Genome-wide association study of placental protein concentrations

Tables 1 and 2 show the characteristics of the 2,352 nulliparous women included in the GWAS of placental protein levels. The heritability estimates from Genome-wide Complex Trait Analysis (GCTA)^54^ indicated that genome-wide SNPs accounted for approximately 14.5% to 31% of the variance in the nine proteins (Supplementary Table 3). The quantile-quantile plots (Supplemental Figure 3) for all GWAS (Supplemental Figure 4) showed no inflation of the test statistics. A summary of all GWAS SNPs with *P<*5×10^-8^ can be found in Supplemental Table 4. The SNP rs6487735, located near the pregnancy zone protein (*PZP*) gene, exhibited the strongest association (MAF=0.47, effect=-0.1, *P=*3.03×10^-22^; Figure 2C) with ADAM-12 levels at visit2. A likely causal missense variant, rs2277413 in the *PZP* gene, independent of rs6487735 (linkage disequilibrium r2=0.003, Figure 2C), was also significantly associated with ADAM-12 levels at visit2 (MAF=0.3, effect=0.07, *P=*3.32×10^-11^) and showed the strongest association after additional adjustment for rs6487735 (*P*=2.37×10^-10^). The GWAS of ADAM-12 at visit1 also identified the same *PZP* locus, which was marginally significant (Figure 2B). The results suggested that *PZP* was associated with ADAM-12 levels in maternal serum, and this association increased as pregnancy progressed. In sensitivity analyses, we repeated the GWAS of ADAM-12 using only those of self-reported as White, most of whom clustered tightly in the PCA plot (Supplemental Figure 1). We obtained similar but less significant results compared to using all available individuals (Supplemental Figure 5 and Supplemental Table 5).

**Table 1.** Summary of women characteristics included in the GWAS of placental protein levels

|  |  | Total (n=2352) |
| --- | --- | --- |
| Age at visit1 (mean±SD) |  | 26.77±5.84 |
| Self-reported race |  |  |
|  | White | 1356 (57.65%) |
|  | Black | 389 (16.54%) |
|  | Hispanic | 394 (16.75%) |
|  | Asian | 80 (3.4%) |
|  | Other | 133 (5.65%) |
| Adverse pregnancy outcome status |  | 1463 (62.2%) |
|  | Preeclampsia or eclampsia | 552 (23.47%) |
|  | Preterm birth | 776 (32.99%) |
|  | Spontaneous preterm birth | 451 (19.18%) |
|  | Stillbirth | 48 (2.04%) |
|  | Small gestational age | 396 (16.84%) |

**Table 2.**
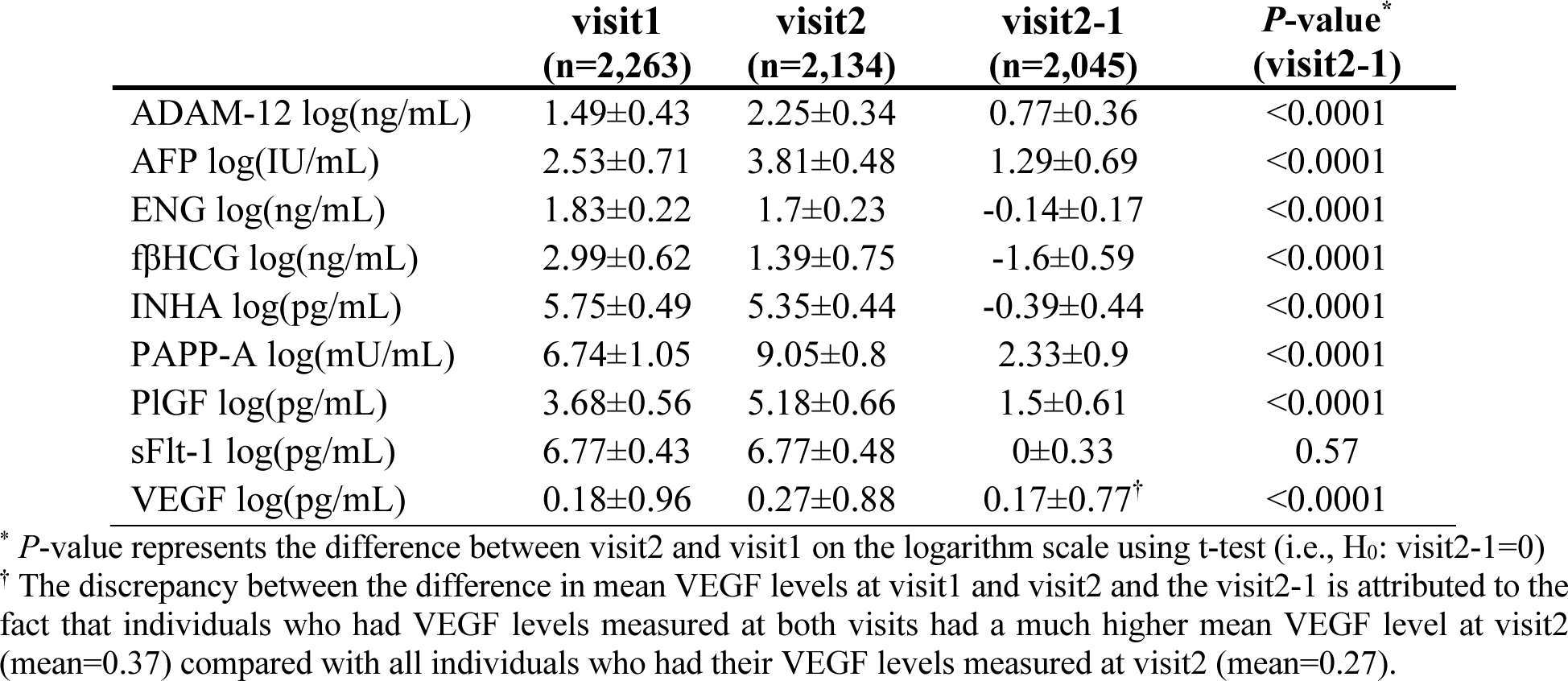
Summary of the serum levels of nine placental proteins (mean±SD) included in GWAS

**Figure 2.**
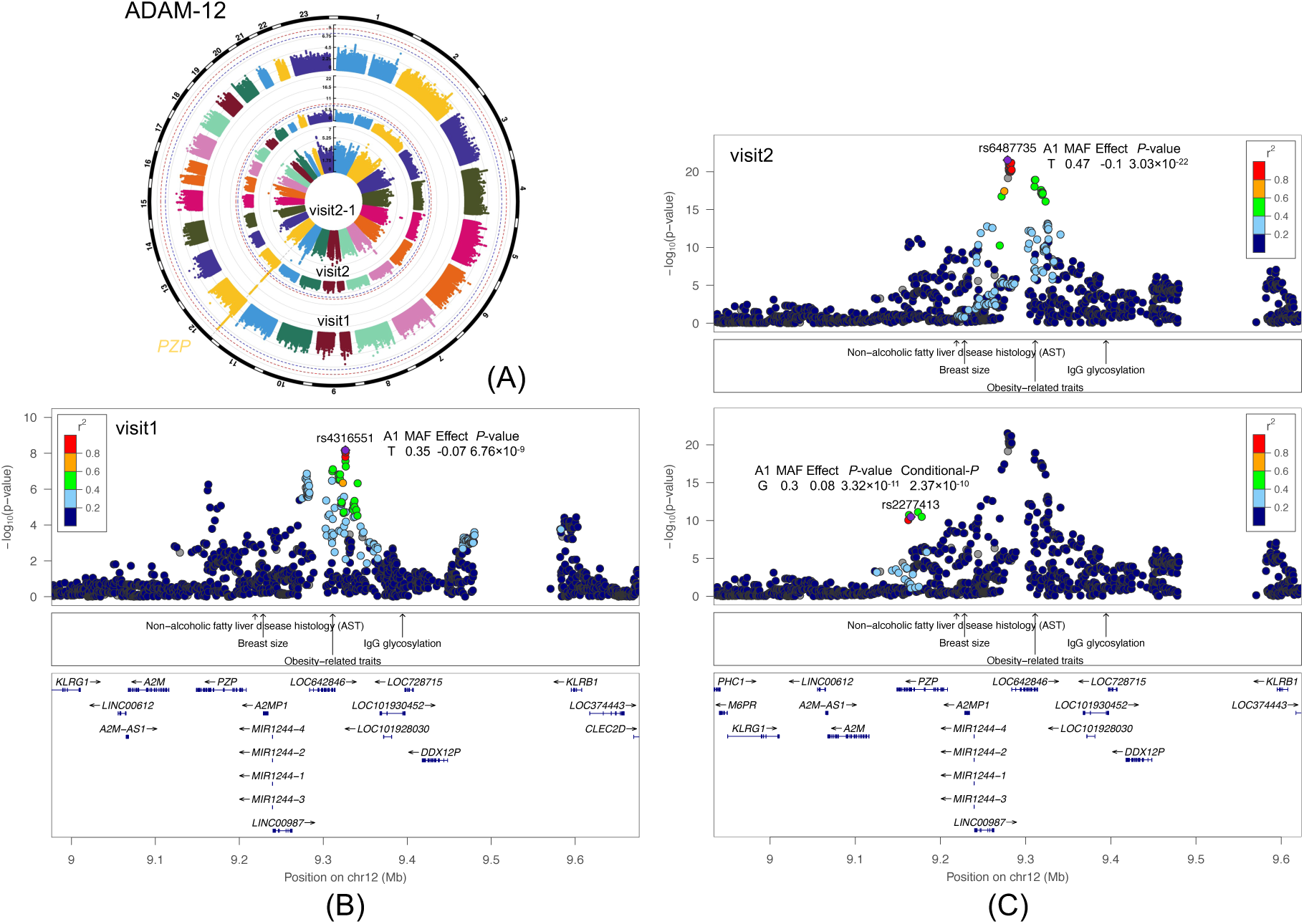
GWAS of ADAM-12. (A) Circular Manhattan plots. Manhattan plot displays the associations between SNPs across the genome and a specific trait, with the spikes indicating regions of significant associations. This circular format presents results from multiple GWAS simultaneously. The chromosomal position of each single SNP is displayed along the circle and the negative log10 of the association *P*-value is displayed on the radius. The red line represents the genome-wide significance level (*P*<5.6×10^-9^) and blue line represents the suggestive significance level (*P*<5×10^-8^). Results for visit1 are displayed on the outer circle, visit2 on the middle circle, and visit2-1 on the inner circle. (B) Regional plot for SNP rs4316551 from the visit1 analysis. A regional plot provides a detailed view of a specific genomic region, showing the association between SNPs and a specific trait. The lower portion of the figure displays the relative location of genes and the direction of transcription, while the middle portion shows known GWAS associations at the locus from the GWAS catalog. The x-axis displays the chromosomal position and the y-axis shows the significance of the associations. The purple diamond shows the *P*-value for the reference SNP. The circles show the *P*-values for all other SNPs and are color coded according to the level of linkage disequilibrium with the reference SNP using the nuMoM2b cohort. (C) Regional plots for SNPs rs6487735 and rs2277413 from the visit2 analysis.

Our GWAS analysis of VEGF identified rs6921438, located near the vascular endothelial growth factor A (*VEGFA*) gene, as the variant with the strongest association with VEGF levels at visit1 (MAF=0.45, effect=-0.36, *P=*7.94×10^-30^) and visit2 (MAF=0.47, effect=-0.32, *P=*2.49×10^-28^) (Figure 3AD). This locus has consistently been replicated in multiple studies as being associated with circulating VEGF levels in non-pregnant populations.^23–26^ Our study confirms the presence of *cis*-acting genetic associations with VEGF levels in nulliparous pregnant women. We subsequently investigated whether SNPs previously linked to VEGF levels in non-pregnant populations at the *VEGFA* and very low-density lipoprotein receptor (*VLDLR*) loci were also linked to VEGF levels in pregnant women. Our analysis confirmed an association with SNPs at *VEGFA*, but not with *VLDLR* (Supplemental Table 6). These findings suggest that the genetic regulation of VEGF levels may differ between early pregnancy and non-pregnant periods.

**Figure 3.**
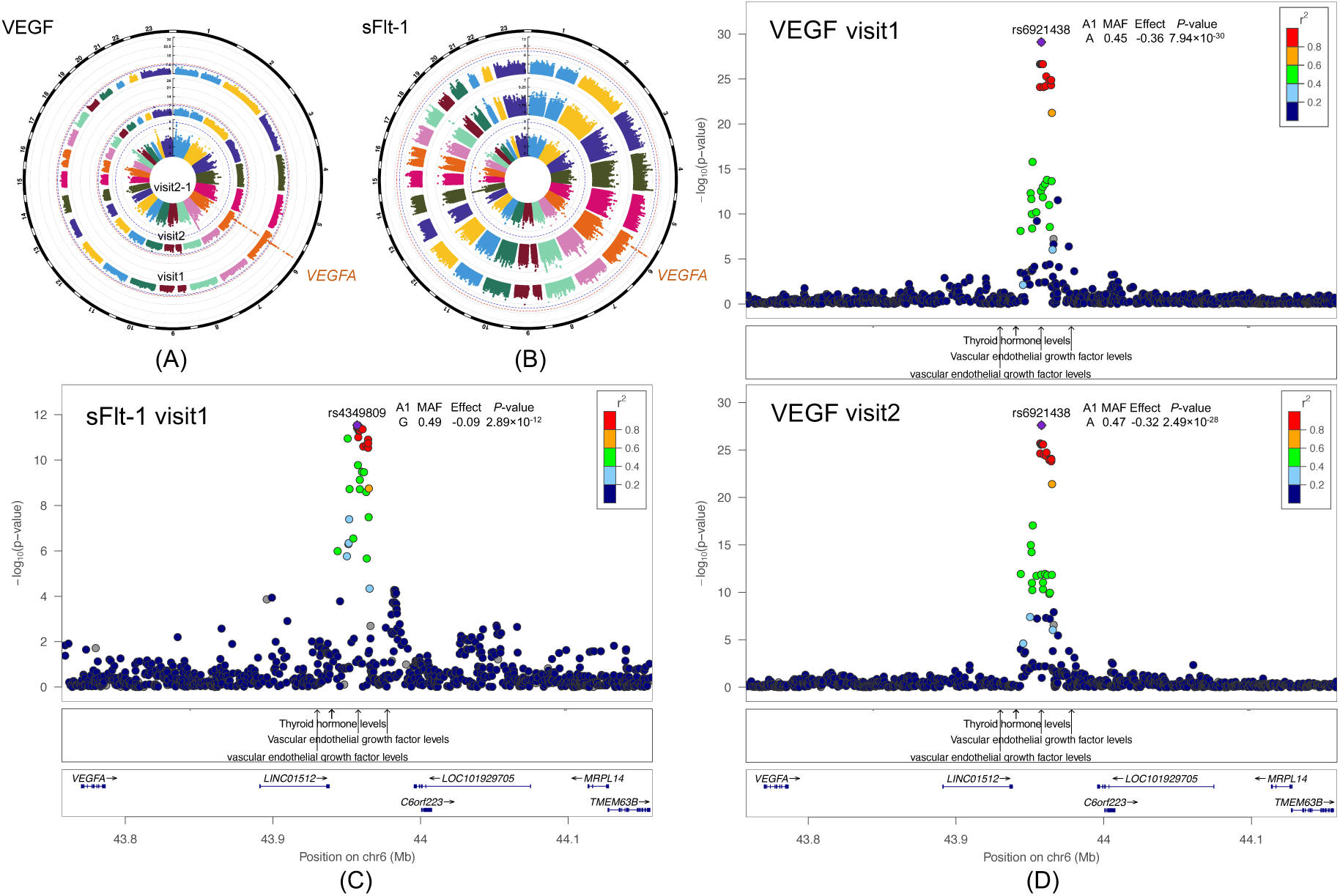
GWAS of sFlt-1 and VEGF. (A) and (B) Circular Manhattan plots for sFlt-1 and VEGF. Manhattan plot displays the associations between SNPs across the genome and a specific trait, with the spikes indicating regions of significant associations. This circular format presents results from multiple GWAS simultaneously. The chromosomal position of each single SNP is displayed along the circle and the negative log10 of the association *P*-value is displayed on the radius. The red line represents the genome-wide significance level (*P*<5.6×10^-9^) and blue line represents the suggestive significance level (*P*<5×10^-8^). Results for visit1 are displayed on the outer circle, visit2 on the middle circle, and visit2-1 on the inner circle. (C) Regional plot for SNP rs4349809 from the sFlt-1 visit1 analysis. A regional plot provides a detailed view of a specific genomic region, showing the association between SNPs and a specific trait. The lower portion of the figure displays the relative location of genes and the direction of transcription, while the middle portion shows known GWAS associations at the locus from the GWAS catalog. The x-axis displays the chromosomal position and the y-axis shows the significance of the associations. The purple diamond shows the *P*-value for the reference SNP. The circles show the *P*-values for all other SNPs and are color coded according to the level of linkage disequilibrium with the reference SNP using the nuMoM2b cohort. (C) Regional plots for SNP rs6921438 from the VEGF visit1 and visit2 analyses.

In the GWAS of sFlt-1, rs4349809 at the same *VEGFA* locus was found to be significantly associated with sFlt-1 levels at visit1 (MAF=0.49, effect=-0.09, *P=*2.89×10^-12^), but this association was not observed at visit2 (Figure 3BC). Furthermore, rs4349809 was also associated with VEGF levels in the same direction at visit1 (effect=-0.33, *P=*7.91×10^-25^) and visit2 (effect=-0.3, *P=*2.34×10^-25^) as with sFlt-1. VEGF stimulates blood vessel growth crucial for supporting fetal development, while sFlt-1 inhibits VEGF by binding to it.^55^ Despite their opposing roles in regulating angiogenesis during pregnancy, both were influenced by the *VEGFA* locus, which showed a stronger association with VEGF than sFlt-1. We performed sensitivity analyses by repeating the GWAS analysis of VEGF and sFlt-1 using only self-reported White. The results were similar to those obtained using all individuals, but less statistically significant (Supplemental Figures 6 and 7 and Supplemental Table 5).

All the significant genetic associations persisted even after further adjustment for gestational age at the time of blood collection (Supplemental Figure 8). To further explore the relationship between VEGF and sFlt-1, we conducted additional GWAS focusing on the VEGF/sFlt-1 ratio at visit1, visit2, and visit2-1. The same *VEGFA* locus was found to be associated with the ratio at visit1 and visit2 (Supplemental Figure 9).

### Evidence for potential causal associations: placental protein levels, preeclampsia/gestational hypertension, and long-term postpartum hypertension

Our MR-RAPS analyses revealed a significant effect between ADAM-12 at visit1 and gHTN (odds ratio (OR)=0.78, *P=*8.6×10^-4^) (Figure 4 and Supplemental Figure 10). Both IVW^51^ and MR-PRESSO^52^ analyses confirmed this association (Supplemental Figures 11 and 12). Although ADAM-12 levels did not meet the significance threshold for a causal link with PE, the consistent direction and similar ORs for ADAM-12 levels at both visits for PE and gHTN (Figure 4) suggest that a similar relationship to PE remains possible. This is consistent with clinical evidence that gHTN is on the spectrum of hypertensive disorders with PE, as it has similar clinical features and frequently progresses to PE when expectantly managed.^56, 57^ Our results also revealed potential causal effects of PE (OR=1.75, *P*=8.3×10^-3^) and gHTN (OR=1.84, *P*=4.7×10^-3^) on the development of long-term postpartum HTN (Figure 5 and Supplemental Figure 13). Having demonstrated the potential causal pathway, ADAM-12 → PE/gHTN and PE/gHTN → long-term postpartum HTN, we conducted further mediation analysis to examine the existence of the ADAM-12 → PE/gHTN → long-term postpartum HTN pathway. The results showed a significant effect, whereby ADAM-12 at visit1, inversely associated with gHTN, was further inversely associated with postpartum HTN 2-7 years later, as estimated by MR-RAPS (mediated effect=-0.15, *P=*0.03). In all Mendelian randomization analyses, we observed no heterogeneity, as indicated by Cochran’s Q, and the MR-PRESSO global pleiotropy test revealed no evidence of horizontal pleiotropy in the IV SNPs.

**Figure 4.**
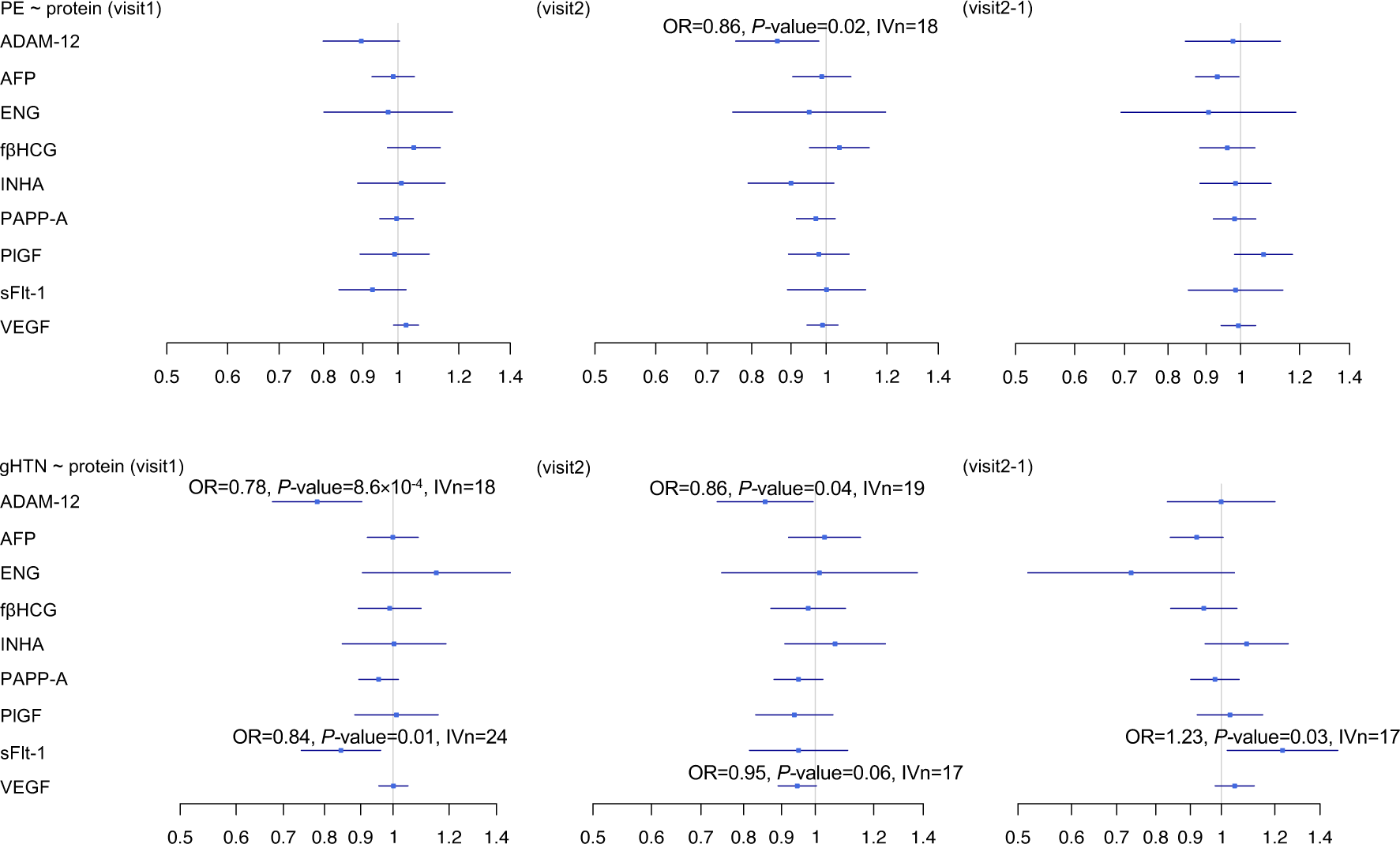
Causal estimates of the serum levels of nine placental proteins on PE and gHTN. *P*-values were determined by the two-sample MR-RAPS method. The squares represent the causal estimates on the odds ratio (OR) scale, and the whiskers show the corresponding 95% confidence intervals.

**Figure 5.**
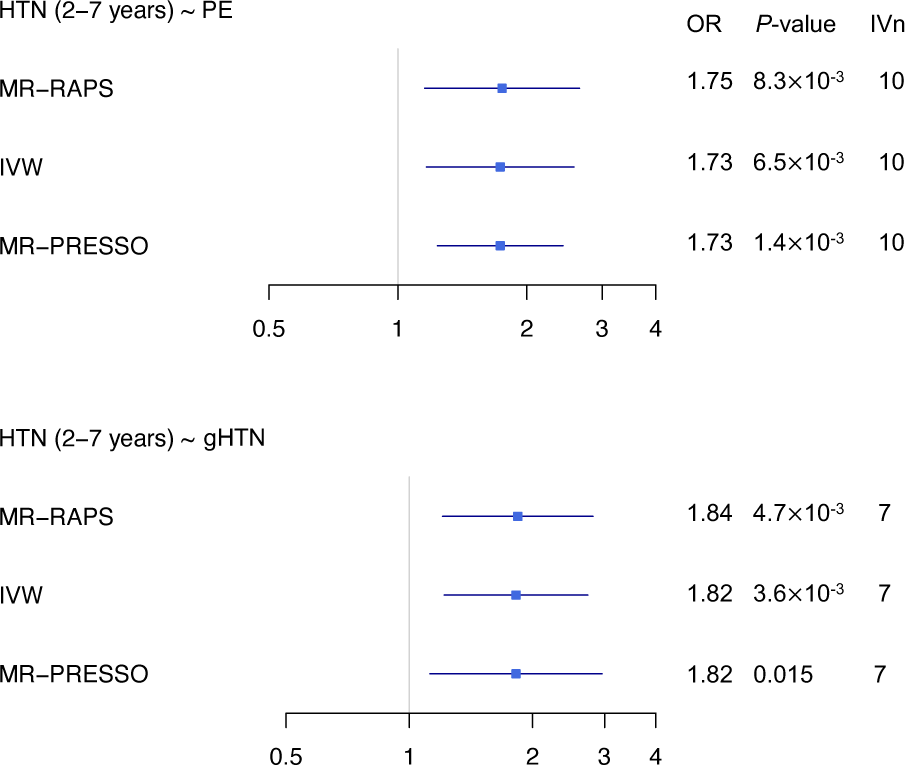
Causal estimates of PE and gHTN on long-term postpartum HTN. *P*-values were determined by the two-sample MR-RAPS, IVW, and MR-PRESSO methods. The squares represent the causal estimates on the odds ratio (OR) scale, and the whiskers show the corresponding 95% confidence intervals.

### Discussion Principal findings

In our GWAS, we identified significant associations between the *PZP* locus and ADAM-12 levels, and the *VEGFA* locus with levels of both VEGF and sFlt-1 during pregnancy. Mendelian randomization analyses provided evidence for a potential causal relationship between ADAM-12 at visit1 and gHTN, as well as between PE/gHTN in the first pregnancy and HTN occurring 2-7 years postpartum.

### Results in the context of what is known

Our identification of *PZP* as being important for ADAM-12 levels is consistent with existing literature on the biological effects of *PZP*. ^58–62^ *PZP* encodes a protein (PZP) that plays a critical protective role during pregnancy by managing inflammation and oxidative stress.^58, 59^ It assists in clearing misfolded proteins and pro-inflammatory cytokines, both enhanced by oxidative stress, to prevent inflammatory responses that could impair placental function.^59^ Additionally, it modulates immune activity by inhibiting T-helper 1 (Th1) cells in conjunction with placental protein-14 (PP14), helping maintain a pregnancy-friendly immune environment and preventing maternal immune rejection of the fetus.^58–60^ Low levels of PZP can lead to uncontrolled inflammation, contributing to placental dysfunction and the onset of conditions like PE.^59^ ADAM-12, a metalloproteinase secreted by the placenta, cleaves insulin-like growth factor binding proteins (IGFBPs).^39, 63^ It promotes cell invasion and direct column outgrowth in early placental development.^62^ ADAM-12 has been localized to anchoring trophoblast columns of first-trimester placentas and to highly invasive trophoblasts within placental villous explants that degrade the extracellular matrix.^61, 62^ Recent studies further highlight the role of its shorter variant, ADAM12S, in regulating the migration and invasion of trophoblasts into the uterine lining.^39^ Moreover, the placenta expresses ADAM-12 at high levels, leading to elevated concentrations in the maternal circulation during pregnancy.^63^ Low first-trimester levels of ADAM-12 in maternal circulation have been consistently associated with the development of PE.^39, 64, 65^ These known functions of ADAM-12 as a key regulator of early placental implantation and development are consistent with our finding of statistical evidence for its a causal role in gHTN and possibly PE.

Our finding that *VEGFA* was associated with VEGF levels at visits 1 and 2 is in line with previous reports in non-pregnant populations.^23–26^ Our study extends these findings to confirm the presence of *cis*-acting genetic associations with VEGF levels during pregnancy. Surprisingly, *VEGFA*’s association with sFlt-1 levels at visit1 showed the same effect direction as for VEGF levels, despite their antagonistic functions during pregnancy.^55^ Both VEGF and sFlt-1 are known to be crucial for the development of the placenta, and their balance is essential for normal placental angiogenesis.^66–68^ VEGF, a potent angiogenic factor, stimulates the formation of new blood vessels and supports endothelial cell function.^66, 67^ sFlt-1, a soluble receptor of VEGF, acts as an anti-angiogenic protein by antagonizing the actions of VEGF and PlGF.^66–68^ An imbalance, characterized by increased levels of sFlt-1 and decreased levels of both VEGF and PlGF, is believed to contribute to the development of PE.^4, 8, 69–73^

### Clinical implications

Our results suggest the potential utility of early pregnancy ADAM-12 serum levels as a biomarker and potential therapeutic target for gHTN and PE. Its established role in early placental development and trophoblast invasion,^61, 62^ a process that is known to be incomplete in PE and other placenta-mediated conditions,^1^ makes it an attractive candidate for early pregnancy disease modification. Finally, our findings suggest a role for ADAM-12 in the long-term development of postpartum HTN, suggesting that early intervention targeting pathways regulated by ADAM-12 may also mitigate the long-term CV risks in women affected by gHTN or PE.

### Research implications

Further work is needed to confirm both the predictive and causal roles of ADAM-12 for hypertensive disorders of pregnancy. ADAM-12 administration and the importance of *PZP* can be investigated in animal models, while confirmation of ADAM-12’s utility as a biomarker requires validation in large human cohorts. Future research should focus on validating the predictive value of ADAM-12, elucidating its biological mechanisms, and exploring aspects of its practical application in clinical settings. While ours was a targeted approach, additional opportunities for biomarker and therapeutic target discovery are available via untargeted proteomics techniques, such as Olink or SomaScan.^74^ Establishing a comprehensive profile of genetically regulated proteins during pregnancy may identify additional biomarkers and therapeutic targets as well as provide a unique resource for further Mendelian randomization studies examining pregnancy and postpartum outcomes. Finally, investigating the fetal genetic influences on placental proteins is essential, as many circulating proteins (e.g., VEGF and sFlt-1^3, 4, 9^) are predominantly derived from the placenta. Future studies incorporating fetal genotypes will also enable researchers to distinguish between genetic contributions originating from the fetus or the mother.

### Strengths and limitations

We analyzed the changes in placental proteins from maternal serum during early pregnancy. These early pregnancy proteins may serve as predictors for outcomes during and after pregnancy. This study is the first to systematically examine multiple protein levels during pregnancy using a GWAS approach. Our findings provide insights into the genetic regulation of placental protein levels during pregnancy. Moreover, we employed the two-sample Mendelian randomization, a method recognized for its conservative and unbiased approach to causal inference,^75^ and took advantage of a recent large PE/gHTN GWAS^29^ in Mendelian randomization analyses.

We recognize several study limitations. First, the sample size was limited, although this is the first GWAS examining the specific phenotypes. Second, while our analytical workflow was designed to minimize bias due to population stratification in our multi-ethnic cohort, it is possible that this approach inadvertently biased the results toward SNPs more common or with stronger effects in White, given their larger proportion. Additionally, this method may have eliminated SNPs with genetic effects specific to a single ancestry. Third, only nine proteins related to placental health were examined. Untargeted proteomics data may identify additional genetically regulated proteins during pregnancy. Fourth, the lack of available data concerning the same questions prevented the replication of the GWAS of placental proteins in other cohorts. Fifth, fetal SNPs, which could be important, are lacking. The use of fetal SNPs as IVs might be a more effective approach in Mendelian randomization analysis to avoid horizontal pleiotropy. Sixth, the correlation between maternal and fetal genotypes makes it challenging to discern the source of the genetic effects.

## Conclusions

In conclusion, our study identified significant genetic associations with placental proteins ADAM-12, VEGF, and sFlt-1, providing insights into their regulation during pregnancy. Mendelian randomization analyses revealed evidence for potential causal relationships between the serum levels of placental proteins, particularly ADAM-12, and PE/gHTN, which could lead to potential prevention and treatment strategies for hypertensive disorders of pregnancy. Further research is needed to understand the biological mechanisms underlying these associations and confirm their causal relationships with PE/gHTN.

## Availability of data and materials

The individual-level data are available through the NICHD’s Data and Specimen Hub (DASH) at https://doi.org/10.57982/gjxm-yz73.

## Conflict of interest

M.C.H. reports consulting fees from CRISPR Therapeutics; advisory board service for Miga Health; and grant support from Genentech (all unrelated to the present work).

## Funding

Support for performing DNA extraction and GWAS from the Indiana University Grand Challenges Precision Diabetes project funding. Q.Y. was partially funded by R03OD034501 from the U.S. National Institutes of Health (NIH). D.M.H. was partially funded by R01HD101246 from the U.S. NIH. M.C.H. was supported by K08HL166687 from the U.S. NIH and American Heart Association (940166, 979465). The nuMoM2b specimen and data collection were supported by Grant funding from the Eunice Kennedy Shriver National Institute of Child Health and Human Development (NICHD): U10HD063036; U10HD063072; U10HD063047; U10HD063037; U10HD063041; U10HD063020; U10HD063046; U10HD063048; and U10HD063053. In addition, support was provided by Clinical and Translational Science Institutes: UL1TR001108 and UL1TR000153. The nuMoM2b Heart Health Study was supported by cooperative agreement funding from the National Heart, Lung, and Blood Institute (NHLBI) and NICHD: U10HL119991; U10HL119989; U10HL120034; U10HL119990; U10HL120006; U10HL119992; U10HL120019; U10HL119993; U10HL120018, and U01HL145358; and the National Center for Advancing Translational Sciences through UL1TR000124, UL1TR000153, UL1TR000439, and UL1TR001108; and the Barbra Streisand Women’s Cardiovascular Research and Education Program, and the Erika J. Glazer Women’s Heart Research Initiative, Cedars-Sinai Medical Center, Los Angeles. The content of this article is solely the responsibility of the authors and does not necessarily represent the official views of NHLBI, NIH, or the US Department of Health and Human Services. The funding sources were not involved in the interpretation of the result or in decisions regarding which journal to submit.

## Data Availability

The individual-level phenotype and genotype data have been reported earlier and are available upon request.

## Glossary of Terms

- **ADAM-12:** A disintegrin and metalloproteinase domain-containing protein 12, involved in cell adhesion, migration, proliferation, and placental function.
- **AFP:** Alpha fetal protein.
- **Bonferroni adjustment:** A method to correct for multiple comparisons in GWAS. It reduces the chance of false positives by lowering the threshold for statistical significance. It is usually set at *P*-value=5×10^-8^ for a single GWAS. When multiple GWAS are performed, further adjustment may be needed.
- **Circular Manhattan plot:** A variant of the traditional Manhattan plot, which is presented in a linear format with chromosomes laid out along the x-axis and the -log10 of the *P*-value on the y-axis. In contrast, a circular Manhattan plot arranges this information in a circular format, allowing multiple Manhattan plots to be displayed within the same circle.
- **Collider bias:** A potential bias in regression models (e.g., Y = X + covariate), where if Y and X independently influence the covariate, collider bias may occur.
- **ENG:** Endoglin.
- **fβHCG:** Free beta-human chorionic gonadotropin.
- **Genome-wide association study (GWAS):** A study assessing the association of SNPs across the genome with a specific trait.
- **Genome-wide interaction studies (GWIS):** A study assessing interactions between SNP-by-exposure and a specific trait across the genome.
- **Genotype:** In the context of GWAS, genotypes are often coded as 0 (homozygous for the reference allele), 1 (heterozygous), or 2 (homozygous for the alternative allele) to reflect the number of alternative alleles carried by an individual.
- **Genotype imputation:** The process of inferring genotypes that are not directly measured.
- **Horizontal pleiotropy:** In Mendelian Randomization, horizontal pleiotropy occurs when instrumental variable SNPs influence the outcome through pathways other than the exposure, as shown in pathway (3) in Figure 1B. It is assumed that horizontal pleiotropy is absent. The existence of horizontal pleiotropy can be assessed by statistical tests, such as Cochran’s Q and the MR-PRESSO global pleiotropy test.
- **INHA:** Inhibin A.
- **Instrumental variables:** In Mendelian Randomization, an instrumental variable is a SNP used as a proxy for the exposure to estimate its causal effect on the outcome.
- **Linear mixed model:** A statistical model that can account for both fixed effects and random effects. In this study, it is used to control for genetic relatedness in association studies.
- **Manhattan plot:** A graphical representation of GWAS results that displays the -log10 of the *P*-value for each SNP across the genome. Each dot represents a single SNP, with its height indicating the level of significance of its association with the outcome. This plot helps to highlight genomic regions significantly associated with the outcome.
- **Mediation analysis:** A statistical method used to understand how the exposure influences the outcome through an intermediate variable, known as a mediator.
- **Mendelian randomization:** A statistical method using SNPs as instrumental variables to estimate the causal effect of a risk factor on an outcome, aiming to minimize unmeasured confounding bias. Multiple approaches are recommended for consistency due to untestable assumptions like horizontal pleiotropy.
- **PAPP-A:** Pregnancy-associated plasma protein A.
- **PlGF:** Placental growth factor.
- **Population stratification:** Confounding in genetic studies due to sampling from different ancestries.
- **Principal component analysis (PCA):** In the context of GWAS, PCA reduces the dimensionality of genetic data, summarizing genetic variations into principal components that represent genetic ancestry.
- ***PZP*:** Genetic locus of pregnancy zone protein, which encodes a protein thought to inhibit T-cell function during pregnancy, thereby helping to prevent fetal rejection.
- **Regional plot:** A detailed visualization of GWAS results focused on a specific genomic region, displaying the association *P*-value relative to genomic positions, local linkage disequilibrium reflecting SNP correlations, and the genes located in the region.
- **sFlt-1:** Soluble fms-like tyrosine kinase-1, a circulating antagonist to VEGF and PlGF.
- **Single nucleotide polymorphism (SNP):** A variation at a single position in the DNA sequence among individuals, which is labeled by the “rs” number, a unique identifier assigned to a specific SNP. The “rs” stands for “reference SNP”.
- **VEGF:** Vascular endothelial growth factor, required for regulating the proliferation, migration, and survival of embryonic endothelial cells during the female reproductive cycle.
- ***VEGFA*:** Genetic locus of vascular endothelial growth factor A, which encodes the VEGF protein.
- ***VLDLR*:** Genetic locus of very low-density lipoprotein receptor.

## Notes

### Author Declarations

Nulliparous women with singleton pregnancies were recruited from hospitals affiliated with eight clinical centers: Case Western University; Columbia University; Indiana University; University of Pittsburgh; Northwestern University; University of California at Irvine; University of Pennsylvania; and University of Utah. The Data Coordinating and Analysis Center is RTI International. Each site's local governing Institutional Review Board(s) approved the protocol and procedures.

### Summary of Updates

To better tailor the manuscript for a clinical audience, we have made significant editorial changes, particularly to the Introduction and Discussion sections. The results remain unchanged since the last version.

## References

1. Jung E, Romero R, Yeo L, et al. The etiology of preeclampsia. Am J Obstet Gynecol 2022;226:S844–S66.

2. Melchiorre K, Giorgione V, Thilaganathan B. The placenta and preeclampsia: villain or victim? Am J Obstet Gynecol 2022;226:S954–S62.

3. Zeisler H, Llurba E, Chantraine F, et al. Predictive Value of the sFlt-1:Plgf Ratio in Women with Suspected Preeclampsia. N Engl J Med 2016;374:13–22.

4. Levine RJ, Maynard SE, Qian C, et al. Circulating angiogenic factors and the risk of preeclampsia. N Engl J Med 2004;350:672–83.

5. Droge LA, Perschel FH, Stutz N, et al. Prediction of Preeclampsia-Related Adverse Outcomes With the sFlt-1 (Soluble fms-Like Tyrosine Kinase 1)/Plgf (Placental Growth Factor)-Ratio in the Clinical Routine: A Real-World Study. Hypertension 2021;77:461–71.

6. Velegrakis A, Kouvidi E, Fragkiadaki P, Sifakis S. Predictive value of the sFlt–1/Plgf ratio in women with suspected preeclampsia: An update (Review). Int J Mol Med 2023;52.

7. Parry S, Carper BA, Grobman WA, et al. Placental protein levels in maternal serum are associated with adverse pregnancy outcomes in nulliparous patients. Am J Obstet Gynecol 2022;227:497 e1–97 e13.

8. Maynard SE, Min JY, Merchan J, et al. Excess placental soluble fms-like tyrosine kinase 1 (sFlt1) may contribute to endothelial dysfunction, hypertension, and proteinuria in preeclampsia. J Clin Invest 2003;111:649–58.

9. Smith GC, Crossley JA, Aitken DA, et al. Circulating angiogenic factors in early pregnancy and the risk of preeclampsia, intrauterine growth restriction, spontaneous preterm birth, and stillbirth. Obstet Gynecol 2007;109:1316–24.

10. Dugoff L, Hobbins JC, Malone FD, et al. First-trimester maternal serum PAPP-A and free-beta subunit human chorionic gonadotropin concentrations and nuchal translucency are associated with obstetric complications: a population-based screening study (the Faster Trial). Am J Obstet Gynecol 2004;191:1446–51.

11. Magnussen EB, Vatten LJ, Smith GD, Romundstad PR. Hypertensive disorders in pregnancy and subsequently measured cardiovascular risk factors. Obstet Gynecol 2009;114:961–70.

12. Bellamy L, Casas JP, Hingorani AD, Williams DJ. Pre-eclampsia and risk of cardiovascular disease and cancer in later life: systematic review and meta-analysis. Bmj 2007;335:974.

13. Vikse BE, Irgens LM, Bostad L, Iversen BM. Adverse perinatal outcome and later kidney biopsy in the mother. J Am Soc Nephrol 2006;17:837–45.

14. Vikse BE, Irgens LM, Leivestad T, Skjaerven R, Iversen BM. Preeclampsia and the risk of end-stage renal disease. N Engl J Med 2008;359:800–9.

15. Tschiderer L, Van Der Schouw YT, Burgess S, et al. Hypertensive disorders of pregnancy and cardiovascular disease risk: a Mendelian randomisation study. Heart 2023.

16. Burgess S, Timpson NJ, Ebrahim S, Davey Smith G. Mendelian randomization: where are we now and where are we going? Int J Epidemiol 2015;44:379–88.

17. Levin MG, Burgess S. Mendelian Randomization as a Tool for Cardiovascular Research: A Review. Jama Cardiol 2024;9:79–89.

18. Bennett DA, Holmes MV. Mendelian randomisation in cardiovascular research: an introduction for clinicians. Heart 2017;103:1400–07.

19. Smith GD, Ebrahim S. Mendelian randomisation at 20 years: how can it avoid hubris, while achieving more? Lancet Diabetes Endocrinol 2024;12:14–17.

20. Botkjaer JA, Borgbo T, Kloverpris S, Noer PR, Oxvig C, Andersen CY. Effect of pregnancy-associated plasma protein-A (PAPP-A) single-nucleotide polymorphisms on the level and activity of PAPP-A and the hormone profile in fluid from normal human small antral follicles. Fertil Steril 2016;106:1778–86 e8.

21. Botkjaer JA, Noer PR, Oxvig C, Andersen CY. Author Correction: A common variant of the pregnancy-associated plasma protein-A (PAPPA) gene encodes a protein with reduced proteolytic activity towards IGF-binding proteins. Sci Rep 2019;9:17523.

22. Ruggiero D, Nutile T, Nappo S, et al. Genetics of Plgf plasma levels highlights a role of its receptors and supports the link between angiogenesis and immunity. Sci Rep 2021;11:16821.

23. AHOLA-Olli AV, Wurtz P, Havulinna AS, et al. Genome-wide Association Study Identifies 27 Loci Influencing Concentrations of Circulating Cytokines and Growth Factors. Am J Hum Genet 2017;100:40–50.

24. Choi SH, Ruggiero D, Sorice R, et al. Six Novel Loci Associated with Circulating Vegf Levels Identified by a Meta-analysis of Genome-Wide Association Studies. PLos Genet 2016;12:e1005874.

25. Debette S, VISVIKIS-Siest S, Chen MH, et al. Identification of cis- and trans-acting genetic variants explaining up to half the variation in circulating vascular endothelial growth factor levels. Circ Res 2011;109:554–63.

26. Sliz E, Kalaoja M, AHOLA-Olli A, et al. Genome-wide association study identifies seven novel loci associating with circulating cytokines and cell adhesion molecules in Finns. J Med Genet 2019;56:607–16.

27. Sun BB, Chiou J, Traylor M, et al. Plasma proteomic associations with genetics and health in the Uk Biobank. Nature 2023;622:329–38.

28. Xu F, Yu EY, Cai X, et al. Genome-wide genotype-serum proteome mapping provides insights into the cross-ancestry differences in cardiometabolic disease susceptibility. Nat Commun 2023;14:896.

29. Honigberg MC, Truong B, Khan RR, et al. Polygenic prediction of preeclampsia and gestational hypertension. Nat Med 2023.

30. Haas DM, Parker CB, Wing DA, et al. A description of the methods of the Nulliparous Pregnancy Outcomes Study: monitoring mothers-to-be (nuMoM2b). Am J Obstet Gynecol 2015;212:539 e1–39 e24.

31. Guerrero RF, Khan RR, Wapner RJ, et al. Genetic Polymorphisms Associated with Adverse Pregnancy Outcomes in Nulliparas. medRxiv 2022.

32. Haas DM, Ehrenthal DB, Koch MA, et al. Pregnancy as a Window to Future Cardiovascular Health: Design and Implementation of the nuMoM2b Heart Health Study. Am J Epidemiol 2016;183:519–30.

33. De Falco S. The discovery of placenta growth factor and its biological activity. Exp Mol Med 2012;44:1–9.

34. Myatt L. Role of placenta in preeclampsia. Endocrine 2002;19:103–11.

35. Wang J, Dong X, Wu HY, et al. Relationship of Liver X Receptors alpha and Endoglin Levels in Serum and Placenta with Preeclampsia. PLos One 2016;11:e0163742.

36. Ciobanu A, Rouvali A, Syngelaki A, Akolekar R, Nicolaides KH. Prediction of small for gestational age neonates: screening by maternal factors, fetal biometry, and biomarkers at 35-37 weeks’ gestation. Am J Obstet Gynecol 2019;220:486 e1–86 e11.

37. Korzeniewski SJ, Romero R, Chaiworapongsa T, et al. Maternal plasma angiogenic index-1 (placental growth factor/soluble vascular endothelial growth factor receptor-1) is a biomarker for the burden of placental lesions consistent with uteroplacental underperfusion: a longitudinal case-cohort study. Am J Obstet Gynecol 2016;214:629 e1–29 e17.

38. Kim M, Park HJ, Seol JW, et al. VEGF-A regulated by progesterone governs uterine angiogenesis and vascular remodelling during pregnancy. Embo Mol Med 2013;5:1415–30.

39. Christians JK, Beristain AG. ADAM12 and PAPP-A: Candidate regulators of trophoblast invasion and first trimester markers of healthy trophoblasts. Cell Adh Migr 2016;10:147–53.

40. Serra B, Mendoza M, Scazzocchio E, et al. A new model for screening for early-onset preeclampsia. Am J Obstet Gynecol 2020;222:608 e1–08 e18.

41. Wald NJ, Watt HC, Hackshaw AK. Integrated screening for Down’s syndrome based on tests performed during the first and second trimesters. N Engl J Med 1999;341:461–7.

42. Jelliffe-Pawlowski LL, Baer RJ, Blumenfeld YJ, et al. Maternal characteristics and mid-pregnancy serum biomarkers as risk factors for subtypes of preterm birth. Bjog 2015;122:1484–93.

43. Brownbill P, Edwards D, Jones C, et al. Mechanisms of alphafetoprotein transfer in the perfused human placental cotyledon from uncomplicated pregnancy. J Clin Invest 1995;96:2220–6.

44. Hughes AE, Sovio U, Gaccioli F, Cook E, CHARNOCK-Jones DS, Smith GCS. The association between first trimester Afp to PAPP-A ratio and placentally-related adverse pregnancy outcome. Placenta 2019;81:25–31.

45. Taliun D, Harris DN, Kessler MD, et al. Sequencing of 53,831 diverse genomes from the Nhlbi TOPMed Program. Nature 2021;590:290–99.

46. Gogarten SM, Sofer T, Chen H, et al. Genetic association testing using the Genesis R/Bioconductor package. Bioinformatics 2019;35:5346–48.

47. Wojcik GL, Graff M, Nishimura KK, et al. Genetic analyses of diverse populations improves discovery for complex traits. Nature 2019;570:514–18.

48. Mcginnis R, Steinthorsdottir V, Williams NO, et al. Variants in the fetal genome near FLT1 are associated with risk of preeclampsia. Nat Genet 2017;49:1255–60.

49. Steinthorsdottir V, Mcginnis R, Williams NO, et al. Genetic predisposition to hypertension is associated with preeclampsia in European and Central Asian women. Nat Commun 2020;11:5976.

50. Zhao Q, Wang J, Hemani G, Bowden J, Small DS. Statistical inference in two-sample summary-data Mendelian randomization using robust adjusted profile score. Ann Statist 2020;48:1742–69.

51. Burgess S, Butterworth A, Thompson SG. Mendelian randomization analysis with multiple genetic variants using summarized data. Genet Epidemiol 2013;37:658–65.

52. Verbanck M, Chen CY, Neale B, Do R. Detection of widespread horizontal pleiotropy in causal relationships inferred from Mendelian randomization between complex traits and diseases. Nat Genet 2018;50:693–98.

53. Carter AR, Sanderson E, Hammerton G, et al. Mendelian randomisation for mediation analysis: current methods and challenges for implementation. Eur J Epidemiol 2021;36:465–78.

54. Yang J, Lee SH, Goddard ME, Visscher PM. GCTA: a tool for genome-wide complex trait analysis. Am J Hum Genet 2011;88:76–82.

55. Shibuya M. Vascular Endothelial Growth Factor (VEGF) and Its Receptor (VEGFR) Signaling in Angiogenesis: A Crucial Target for Anti- and Pro-Angiogenic Therapies. Genes Cancer 2011;2:1097–105.

56. Acog Practice Bulletin No. 202: Gestational Hypertension and Preeclampsia. Obstet Gynecol 2019;133:1.

57. Koopmans CM, Bijlenga D, Groen H, et al. Induction of labour versus expectant monitoring for gestational hypertension or mild pre-eclampsia after 36 weeks’ gestation (HYPITAT): a multicentre, open-label randomised controlled trial. Lancet 2009;374:979–88.

58. Oh JW, Kim SK, Cho KC, et al. Proteomic analysis of human follicular fluid in poor ovarian responders during in vitro fertilization. Proteomics 2017;17.

59. Wyatt AR, Cater JH, Ranson M. Pzp and PAI-2: Structurally-diverse, functionally similar pregnancy proteins? Int J Biochem Cell Biol 2016;79:113–17.

60. Skornicka EL, Kiyatkina N, Weber MC, Tykocinski ML, Koo PH. Pregnancy zone protein is a carrier and modulator of placental protein-14 in T-cell growth and cytokine production. Cell Immunol 2004;232:144–56.

61. Biadasiewicz K, Fock V, Dekan S, et al. Extravillous trophoblast-associated ADAM12 exerts pro-invasive properties, including induction of integrin beta 1-mediated cellular spreading. Biol Reprod 2014;90:101.

62. Aghababaei M, Perdu S, Irvine K, Beristain AG. A disintegrin and metalloproteinase 12 (ADAM12) localizes to invasive trophoblast, promotes cell invasion and directs column outgrowth in early placental development. Mol Hum Reprod 2014;20:235–49.

63. Loechel F, Fox JW, Murphy G, Albrechtsen R, Wewer UM. Adam 12-S cleaves IGFBP-3 and IGFBP-5 and is inhibited by TIMP-3. Biochem Biophys Res Commun 2000;278:511–5.

64. Laigaard J, Sorensen T, Placing S, et al. Reduction of the disintegrin and metalloprotease ADAM12 in preeclampsia. Obstet Gynecol 2005;106:144–9.

65. Spencer K, Cowans NJ, Stamatopoulou A. ADAM12s in maternal serum as a potential marker of pre-eclampsia. Prenat Diagn 2008;28:212–6.

66. Bolatai A, He Y, Wu N. Vascular endothelial growth factor and its receptors regulation in gestational diabetes mellitus and eclampsia. J Transl Med 2022;20:400.

67. Eddy AC, Bidwell GL, 3RD, George EM. Pro-angiogenic therapeutics for preeclampsia. Biol Sex Differ 2018;9:36.

68. Palmer KR, Tong S, Kaitu’u-Lino TJ. Placental-specific sFLT-1: role in pre-eclamptic pathophysiology and its translational possibilities for clinical prediction and diagnosis. Mol Hum Reprod 2017;23:69–78.

69. Nagamatsu T, Fujii T, Kusumi M, et al. Cytotrophoblasts up-regulate soluble fms-like tyrosine kinase-1 expression under reduced oxygen: an implication for the placental vascular development and the pathophysiology of preeclampsia. Endocrinology 2004;145:4838–45.

70. Shibata E, Rajakumar A, Powers RW, et al. Soluble fms-like tyrosine kinase 1 is increased in preeclampsia but not in normotensive pregnancies with small-for-gestational-age neonates: relationship to circulating placental growth factor. J Clin Endocrinol Metab 2005;90:4895–903.

71. Levine RJ, Qian C, Maynard SE, Yu KF, Epstein FH, Karumanchi SA. Serum sFlt1 concentration during preeclampsia and mid trimester blood pressure in healthy nulliparous women. Am J Obstet Gynecol 2006;194:1034–41.

72. Palmer KR, Kaitu’u-Lino TJ, Hastie R, et al. Placental-Specific sFLT-1 e15a Protein Is Increased in Preeclampsia, Antagonizes Vascular Endothelial Growth Factor Signaling, and Has Antiangiogenic Activity. Hypertension 2015;66:1251–9.

73. Souders CA, Maynard SE, Yan J, et al. Circulating Levels of sFlt1 Splice Variants as Predictive Markers for the Development of Preeclampsia. Int J Mol Sci 2015;16:12436–53.

74. Sun BB, Maranville JC, Peters JE, et al. Genomic atlas of the human plasma proteome. Nature 2018;558:73–79.

75. Burgess S, Davey Smith G, Davies NM, et al. Guidelines for performing Mendelian randomization investigations. Wellcome Open Res 2019;4:186.

