## Supplementary text, figures and tables for "Genetic Associations with Placental Proteins in Maternal Serum Identify Biomarkers for Hypertension in Pregnancy"

### Measurement of placental protein levels

During three study visits in pregnancy, which occurred at 6-13 weeks, 16-21 weeks, and 22-29 weeks of gestation, peripheral maternal blood samples were collected using serum separating tubes. The collected blood samples were then centrifuged, and 0.5 cc serum aliquots were stored at -70  C within two hours of collection. The samples were transported on dry ice to analytical laboratories for batch analyses. Placental protein levels in maternal serum samples collected at the first two study visits were measured for adverse pregnancy outcome (APO) prevention studies as earlier pregnancy biomarkers may provide effective strategies for preventing APOs.

Maternal serum levels of placental proteins were measured at two laboratories: Translational Core Laboratory at Children's Hospital of Philadelphia (Philadelphia, PA) and Eurofins NTD, LLC (Melville, NY). The Children's Hospital laboratory used enzyme-linked immunosorbent assays (ELISA) to measure ENG (human endoglin assay; R&D Systems, Minneapolis, MN) and ADAM-12 (human ADAM-12 ELISA; R&D Systems, Minneapolis, MN), and electrochemiluminescence assays (ECL) to measure VEGF (human VEGF-A ECL; Merck Sharp & Dohme, Kenilworth, NJ) and sFlt-1 (human Flt-1 ECL; Merck Sharp & Dohme, Kenilworth, NJ). The Eurofins NTD laboratory used lanthanide-based time-resolved fluorometry (TRF) to measure five proteins: PlGF, PAPP-A, INHA, fβHCG, and AFP. For more details, please refer to our previous paper.[1]

### Analytical workflow for quality control (QC) and GWAS with multi-ethnic data

#### Genotype data QC and imputation

To conduct pre-imputation QC, we initially excluded SNPs and women with a missing rate greater than 10% in the nuMoM2b cohort. Subsequently, we identified SNPs with Hardy-Weinberg equilibrium (HWE) $P < 1\times10^{-6}$ or minor allele frequency (MAF) < 0.01 in self-reported White, Black, or Hispanic populations, which constitute the three major ethnic groups. These SNPs were then removed from the full dataset. We subsequently performed principal component analysis (PCA) using PC-AiR[2] to account for related individuals, with kinship coefficients estimated by KING.[3] Next, we assessed sex concordance and autosomal heterozygosity, accounting for population structure with the first six principal components (PCs), following the approach described in Bycroft et al. S3.5.2.[4] In brief, we fitted the following linear regression model for the raw sex concordance or autosomal heterozygosity value, $F$,

$$F = F_0 + \sum_{i=1}^{6} \beta_i PC_i + \sum_{i=1}^{6} \sum_{j=i}^{6} \beta_{ij} PC_i PC_j + \varepsilon$$

where the fitted $F_0$ is the PC-adjusted (i.e., ancestry-corrected) sex concordance or autosomal heterozygosity F-value. We then excluded women from the data who violated sex concordance (adjusted F-value > 0.2) or autosomal heterozygosity (adjusted |F-value| > 0.15) (Supplemental Figure 14). As a result, a few individuals were removed from the data. We then repeated the previously mentioned steps for HWE and MAF filters and recalculated the PCA (Supplemental Figure 1).

Next, we used the TOPMed Imputation Server[5] to perform genotype imputation, using the TOPMed (Version R2 on GRC38) as the reference panel, which includes all populations in our cohort. Joint imputation was applied to all individuals, as it is expected to perform as well as splitting the reference panel to match the target population.[6] We retained genotyped and imputed SNPs with imputation quality $r^2 > 0.3$ and repeated HWE and MAF filters. Following QC, our nuMoM2b cohort comprised 9,742 women. For protein genome-wide association studies (GWAS), we analyzed 2,263, 2,134, and 2,045 women for visit1, visit2, and visit2-1 analyses, respectively. Due to our moderate sample size, we focused on SNPs with MAF > 0.05.

GWAS adjusting for population structure

To account for population stratification and genetic relatedness among individuals, we employed a mixed-model approach to conduct GWAS of protein levels. This approach is advantageous because it includes all individuals, regardless of familial and ancestral relatedness, and can provide greater statistical power by controlling for the variance attributed to genetic relatedness.[7] We used the GENESIS R/Bioconductor package[8, 9] to fit linear mixed models that integrated a random effect to control for genetic relatedness, with the genetic relationship matrix (GRM) computed by PC-Relate.[10] The covariates considered in the analysis included age and age-squared at visit1, first 10 PCs calculated by genotypic data, self-reported race, clinical sites, and status for any APOs. To model the continuous log-transformed levels of each protein, $y$, we used a linear mixed model:

$$y = \beta_0 + \beta_1 \text{SNP} + \boldsymbol{\beta}_{\text{cov}} \textbf{covariates} + u + \varepsilon$$

where $u$ is the random variable accounting for genetic relatedness, and $\varepsilon$ is the random error. Although mixed models are advantageous for controlling population stratification, their effectiveness may be limited in a diverse cohort. This is because the GRM, which is estimated based on genome-wide data, assumes that all SNPs have similar population-level deviations. However, some SNPs may have larger or smaller deviations between populations than the genome-wide average, leading to inadequate control of false positive rates.[11] As a result, further adjustments are necessary to address this issue.

Hence, we conducted supplementary genome-wide interaction studies (GWIS) to explore the association between SNP-by-PCs and protein levels,

$$y = \beta_0 + \beta_1 \text{SNP} + \boldsymbol{\beta}_{\text{int}} \text{SNP} \times \textbf{PCs} + \boldsymbol{\beta}_{\text{cov}} \textbf{covariates} + u + \varepsilon$$

where $\textbf{PCs}$ are the first 10 PCs that capture the ancestry information and thus $\boldsymbol{\beta}_{\text{int}}$ includes 10 parameters. We then used a 10 degree of freedom test to evaluate the null hypothesis $H_0$: $\boldsymbol{\beta}_{\text{int}} = 0$. A significant $\boldsymbol{\beta}_{\text{int}}$ indicates that the effects of a particular SNP differ across ancestries. The GWIS were also performed using the GENESIS package.[8, 9] To ensure the retention of only SNPs with consistent effects, we removed SNPs with a *P*-value of $\boldsymbol{\beta}_{\text{int}} < 0.01$ from the GWAS results. It is worth noting that the threshold of 0.01 is conservative and excludes any potentially ancestry-specific SNPs.

Collider bias consideration

The GWAS of protein levels were based on a combination of all women with APOs and a random subset. As a result, the protein levels might not accurately reflect the distributions in the general population. Although women in the nuMoM2b cohort had not progressed to APOs during visit1 and visit2, we adjusted for the APO status in our GWAS of protein levels. However, if both the protein and SNP are independent causes of APO status, there is a possibility of collider bias (as illustrated in Supplemental Figure 15). To address this issue, we performed a GWAS of APO status and examined the association between protein levels and APO status, while adjusting for the same covariates as before except for the APO status, which was the binary outcome in this analysis. We identified the protein-SNP pair as a potential collider bias and excluded it when the *P*-value was less than 0.01 in the GWAS of APO status and when the *P*-value of the association between the protein and APO status was less than 0.05.

Using the proposed GWAS pipeline, we examined genetic associations with the serum levels of nine placental proteins, which were measured during visit1, visit2, and visit2-1. We set the genome-wide significance level at $P < 5.6\times10^{-9}$ (Bonferroni-adjusted for nine proteins: $5\times10^{-8}/9 = 5.6\times10^{-9}$) to identify significant SNPs, which were annotated using ANNOVAR.[12] Additionally, we performed sensitivity analyses by restricting the GWAS to self-reported White only. We conducted the analyses using PLINK2[13] without considering a random effect that controls for genetic relatedness.

**Causal Inference**

Figure 3 illustrates the design of the Mendelian randomization study for causal inference. To investigate the causal relationships between protein levels and both preeclampsia (PE) and gestational hypertension (gHTN), as well as PE/gHTN and long-term postpartum HTN, we used a two-sample Mendelian randomization framework. We also conducted a causal mediation analysis for proteins → PE/gHTN → long-term postpartum HTN. Two-sample Mendelian randomization has a major advantage over one-sample Mendelian randomization as it only requires GWAS summary statistics, rather than individual-level data. Additionally, two-sample Mendelian randomization is typically considered more conservative and unbiased than one-sample Mendelian randomization because it allows for separate cohorts for exposure and outcome data, while in one-sample Mendelian randomization, both exposure and outcome are from the same cohort.[14]

Proteins → PE/gHTN

We used independent SNPs with a $P < 1\times10^{-5}$ from the GWAS of protein levels as instrumental variables (IVs). Our rationale for using this threshold was to identify more independent IV SNPs, as this could promote balanced pleiotropy, which helps mitigate bias due to horizontal pleiotropy. Previous studies have shown that this liberal threshold can result in better performance than a conservative threshold of $5\times10^{-8}$.[15] To select independent IV SNPs, we used a stepwise selection strategy for each chromosome. This approach involved: (1) selecting SNPs with a GWAS $P < 1\times10^{-5}$, ordering them by $P$-values, and selecting the top SNP with the lowest $P$-value; (2) running the genetic regression models again on the remaining SNPs, with additional adjustment on the saved top SNP; (3) selecting SNPs with a conditional $P < 1\times10^{-5}$, ordering them by $P$-values, and saving the top conditional SNP; (4) rerunning the genetic regression models again on the remaining

SNPs, with additional adjustment on the last saved top conditional SNP; and (5) repeating steps (3) and (4) until no SNPs remained in the remaining set.

We obtained genetic association estimates for PE and gHTN from a recent multi-ancestry meta-analysis of GWAS, which included 17,150 PE cases and 451,241 controls, and 8,961 gHTN cases and 184,925 controls in the discovery analysis. It's important to note that the nuMoM2b cohort was not included in the discovery analysis, but rather treated as a follow-up cohort in that study. A detailed description of the study design and participant characteristics can be found in the original publication.[16] After extracting the genetic effect estimates from the GWAS of PE/gHTN, we switched the effect directions and test alleles to ensure consistency with the results from the GWAS of protein levels. Ideally, both studies in a two-sample Mendelian randomization should include the same populations, as some SNPs are expected to have ancestry-specific effects. However, since we excluded SNPs from the GWAS of protein levels that had ancestry-specific effects on protein levels, the genetic results were considered to be generic, and therefore, there were fewer concerns about population compatibility with a second study when conducting a two-sample Mendelian randomization in this current study.

We used MR-robust adjusted profile scoring (MR-RAPS)[17] as the primary method, with squared error loss, as it can account for weak instrument bias which is particularly relevant in our study with a limited number of strong IV SNPs. We also used the commonly used Mendelian randomization methods, including random-effect inverse variance weighting (IVW),[18] and Mendelian randomization pleiotropy residual sum and outlier (MR-PRESSO),[19] which corrects pleiotropy via outlier IV removal. However, we did not use Mendelian randomization-Egger (MR-Egger) in this study, as it is a conservative method,[20] especially when only a few genetic loci are associated with placental protein levels in our GWAS. The reason for using these different methods is that they make different assumptions, and if they produce similar effect estimates, this provides greater confidence in any causal claims. We also assessed the robustness of our results by conducting analyses to identify potential violations of Mendelian randomization assumptions, including heterogeneity measured by Cochran's Q statistic for IVW analyses[18] and horizontal pleiotropy measured by the MR-PRESSO global pleiotropy test.[19] To ensure statistical significance, we set the threshold at $P < 5.6\times10^{-3}$ for the primary analysis, Bonferroni-adjusted for nine proteins ($0.05/9 = 5.6\times10^{-3}$).

PE/gHTN → long-term postpartum HTN

We used SNPs with a $P < 5\times10^{-8}$ from a recent multi-ancestry meta-analysis of GWAS of PE/gHTN as IVs. To investigate genetic associations with long-term postpartum HTN, we conducted a GWAS of HTN occurring 2-7 years after the first pregnancy (972 cases and 3,409 controls) using the nuMoM2b-HHS cohort according to the proposed GWAS pipeline. Long-term HTN was defined as SBP/DBP ≥ 130/80 mmHg or use of antihypertensive medication 2-7 years after the first pregnancy. Subsequently, we conducted a two-sample Mendelian randomization analysis using MR-RAPS, IVW, and MR-PRESSO to assess the causal relationships between PE/gHTN during the first pregnancy and long-term postpartum HTN.

Proteins → PE/gHTN → long-term HTN

The two Mendelian randomization analyses we performed, one for proteins → PE/gHTN and another for PE/gHTN → long-term postpartum HTN, can be used for mediation analysis.[21] We used the product of coefficients method to estimate the mediated effect ($\alpha\beta$), where $\alpha$ is the effect of protein levels → PE/gHTN and $\beta$ is the effect of PE/gHTN → long-term postpartum HTN. To test the composite null hypothesis $H_0: \alpha\beta = 0$, we estimated the standard error of $\alpha\beta$ using the multivariate delta method based on a first-order Taylor series approximation, $\sigma_{\alpha\beta} = \sqrt{\alpha^2\sigma_\beta^2 + \beta^2\sigma_\alpha^2}$.[22] This analysis was conducted solely on ADAM-12 at visit1 due to the strong association it showed with PE/gHTN.

**Supplemental References**

1. Parry S, Carper BA, Grobman WA, et al. Placental protein levels in maternal serum are associated with adverse pregnancy outcomes in nulliparous patients. Am J Obstet Gynecol 2022;227:497 e1-97 e13.
2. Conomos MP, Miller MB, Thornton TA. Robust inference of population structure for ancestry prediction and correction of stratification in the presence of relatedness. Genet Epidemiol 2015;39:276-93.
3. Manichaikul A, Mychaleckyj JC, Rich SS, Daly K, Sale M, Chen WM. Robust relationship inference in genome-wide association studies. Bioinformatics 2010;26:2867-73.
4. Bycroft C, Freeman C, Petkova D, et al. The UK Biobank resource with deep phenotyping and genomic data. Nature 2018;562:203-09.
5. Taliun D, Harris DN, Kessler MD, et al. Sequencing of 53,831 diverse genomes from the NHLBI TOPMed Program. Nature 2021;590:290-99.
6. Howie B, Fuchsberger C, Stephens M, Marchini J, Abecasis GR. Fast and accurate genotype imputation in genome-wide association studies through pre-phasing. Nat Genet 2012;44:955-9.
7. Peterson RE, Kuchenbaecker K, Walters RK, et al. Genome-wide Association Studies in Ancestrally Diverse Populations: Opportunities, Methods, Pitfalls, and Recommendations. Cell 2019;179:589-603.
8. Gogarten SM, Sofer T, Chen H, et al. Genetic association testing using the GENESIS R/Bioconductor package. Bioinformatics 2019;35:5346-48.
9. Wojcik GL, Graff M, Nishimura KK, et al. Genetic analyses of diverse populations improves discovery for complex traits. Nature 2019;570:514-18.
10. Conomos MP, Reiner AP, Weir BS, Thornton TA. Model-free Estimation of Recent Genetic Relatedness. Am J Hum Genet 2016;98:127-48.
11. Conomos MP, Reiner AP, McPeek MS, Thornton TA. Genome-wide control of population structure and relatedness in genetic association studies via linear mixed models with orthogonally partitioned structure. bioRxiv 2018.
12. Wang K, Li M, Hakonarson H. ANNOVAR: functional annotation of genetic variants from high-throughput sequencing data. Nucleic Acids Res 2010;38:e164.
13. Chang CC, Chow CC, Tellier LC, Vattikuti S, Purcell SM, Lee JJ. Second-generation PLINK: rising to the challenge of larger and richer datasets. Gigascience 2015;4:7.
14. Burgess S, Davey Smith G, Davies NM, et al. Guidelines for performing Mendelian randomization investigations. Wellcome Open Res 2019;4:186.
15. Dudbridge F. Power and predictive accuracy of polygenic risk scores. PLoS Genet 2013;9:e1003348.
16. Honigberg MC, Truong B, Khan RR, et al. Polygenic prediction of preeclampsia and gestational hypertension. Nat Med 2023;29:1540-49.
17. Zhao Q, Wang J, Hemani G, Bowden J, Small DS. Statistical inference in two-sample summary-data Mendelian randomization using robust adjusted profile score. Ann Statist 2020;48:1742-69.
18. Burgess S, Butterworth A, Thompson SG. Mendelian randomization analysis with multiple genetic variants using summarized data. Genet Epidemiol 2013;37:658-65.

19. Verbanck M, Chen CY, Neale B, Do R. Detection of widespread horizontal pleiotropy in causal relationships inferred from Mendelian randomization between complex traits and diseases. Nat Genet 2018;50:693-98.
20. Bowden J, Davey Smith G, Burgess S. Mendelian randomization with invalid instruments: effect estimation and bias detection through Egger regression. Int J Epidemiol 2015;44:512-25.
21. Carter AR, Sanderson E, Hammerton G, et al. Mendelian randomisation for mediation analysis: current methods and challenges for implementation. Eur J Epidemiol 2021;36:465-78.
22. MacKinnon DP, Lockwood CM, Hoffman JM, West SG, Sheets V. A comparison of methods to test mediation and other intervening variable effects. Psychol Methods 2002;7:83-104.

**Supplemental Figure 1**

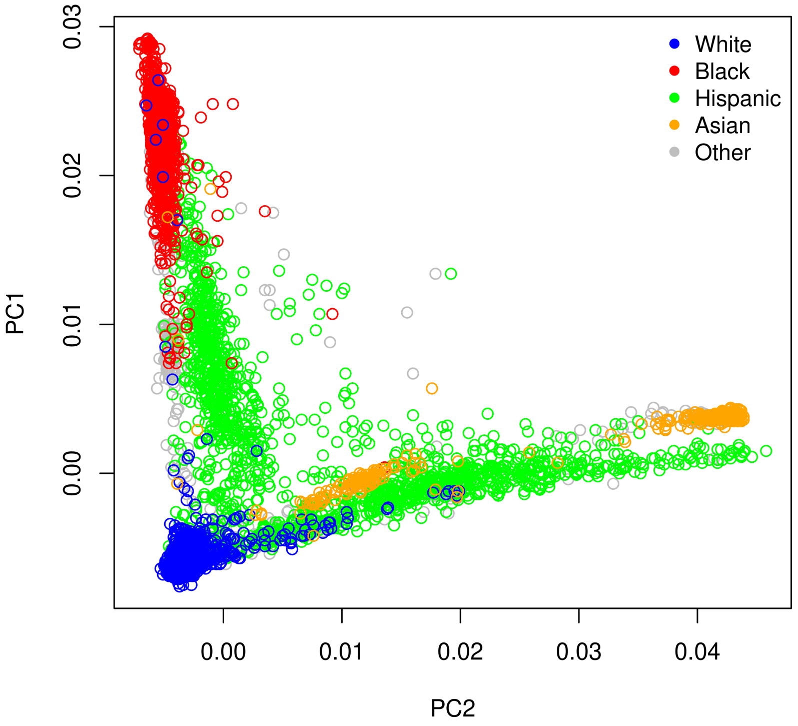

**The first two principal components (PC) from PCA using the nuMoM2b cohort.** Each dot represents an individual, and the dots are color-coded according to self-reported race.

**Supplemental Figure 2**

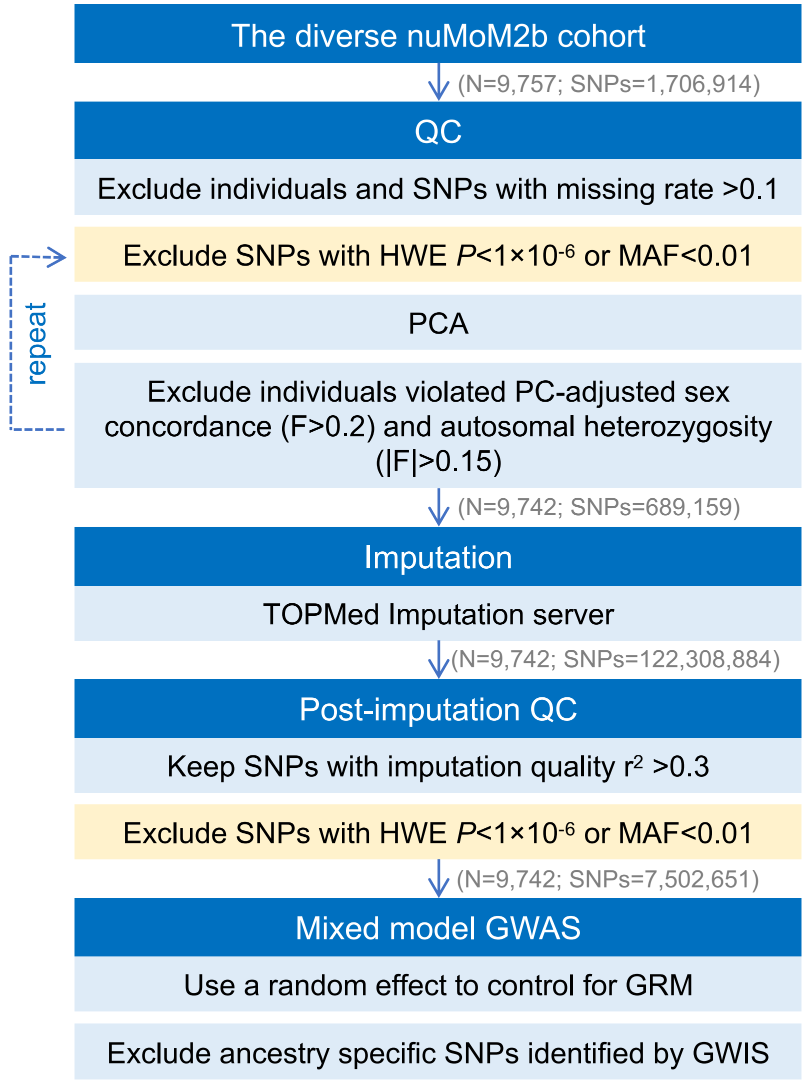

**Flowchart for quality control (QC), genotype imputation, and association test in the diverse nuMoM2b cohort.** The light blue block indicates that the step is conducted on the full diverse cohort, while the light orange block indicates that the step is conducted separately for White, Black, and Hispanic.

### Supplemental Figure 3

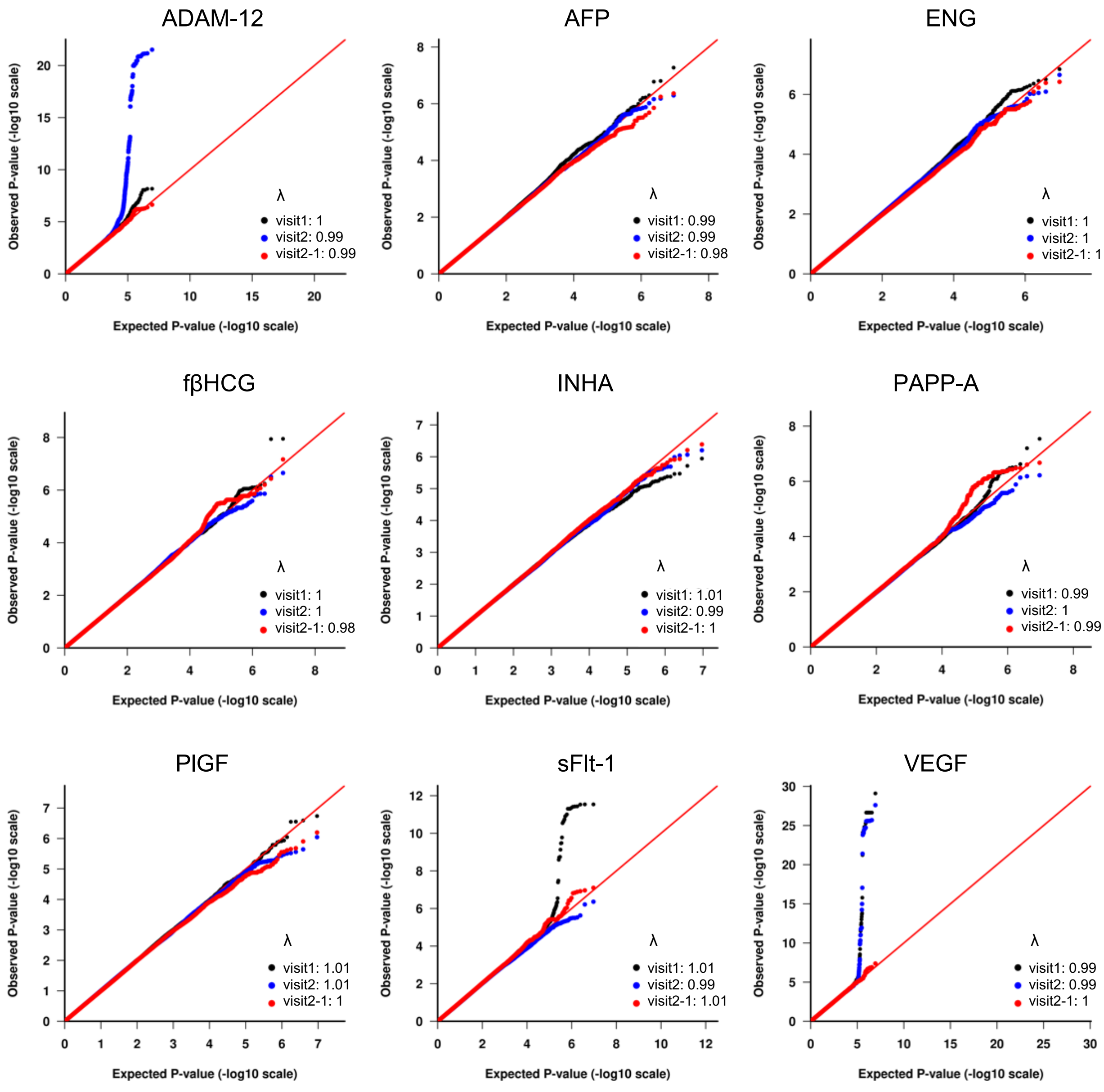

**The quantile-quantile plots.** λ is the genomic control value. The plot displays the distribution of *P*-values against a theoretical distribution, under the null hypothesis of no association. Points (representing SNPs) that align with the diagonal line suggest adherence to the null hypothesis, with a genomic control value near one indicating no inflation. Upward deviations from this line at the higher end indicate SNPs significantly associated with the trait beyond random chance.

**Supplemental Figure 4**

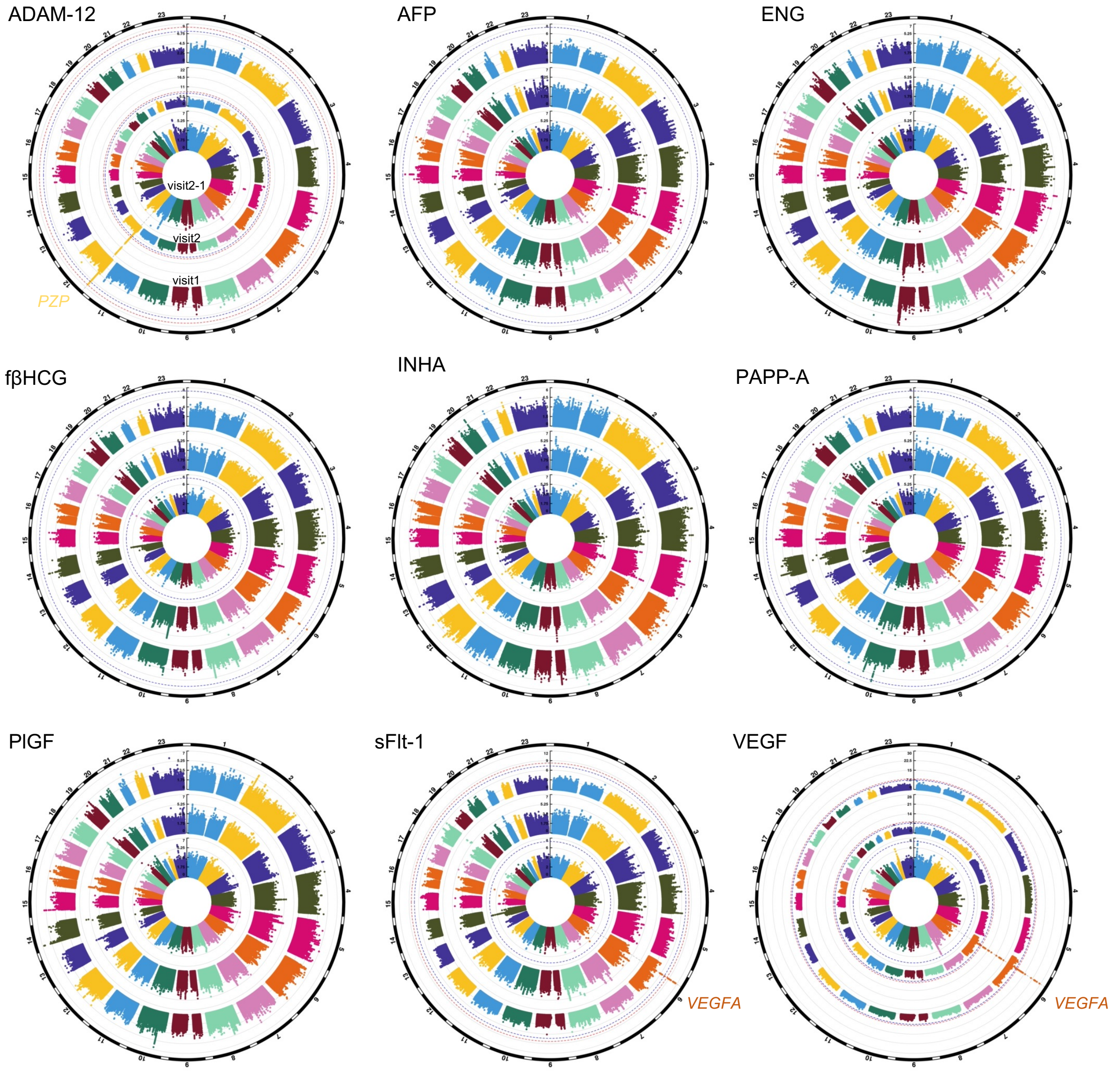

**Circular Manhattan plots.** Manhattan plot displays the associations between SNPs across the genome and a specific trait, with the spikes indicating regions of significant associations. This circular format presents results from multiple GWAS simultaneously. The chromosomal position of each single SNP is displayed along the circle and the negative log10 of the association *P*-value is displayed on the radius. The red line represents the genome-wide significance level ($P < 5.6\times10^{-9}$) and blue line represents the suggestive significance level ($P < 5\times10^{-8}$). Results for visit1 are displayed on the outer circle, visit2 on the middle circle, and visit2-1 on the inner circle.

### Supplemental Figure 5

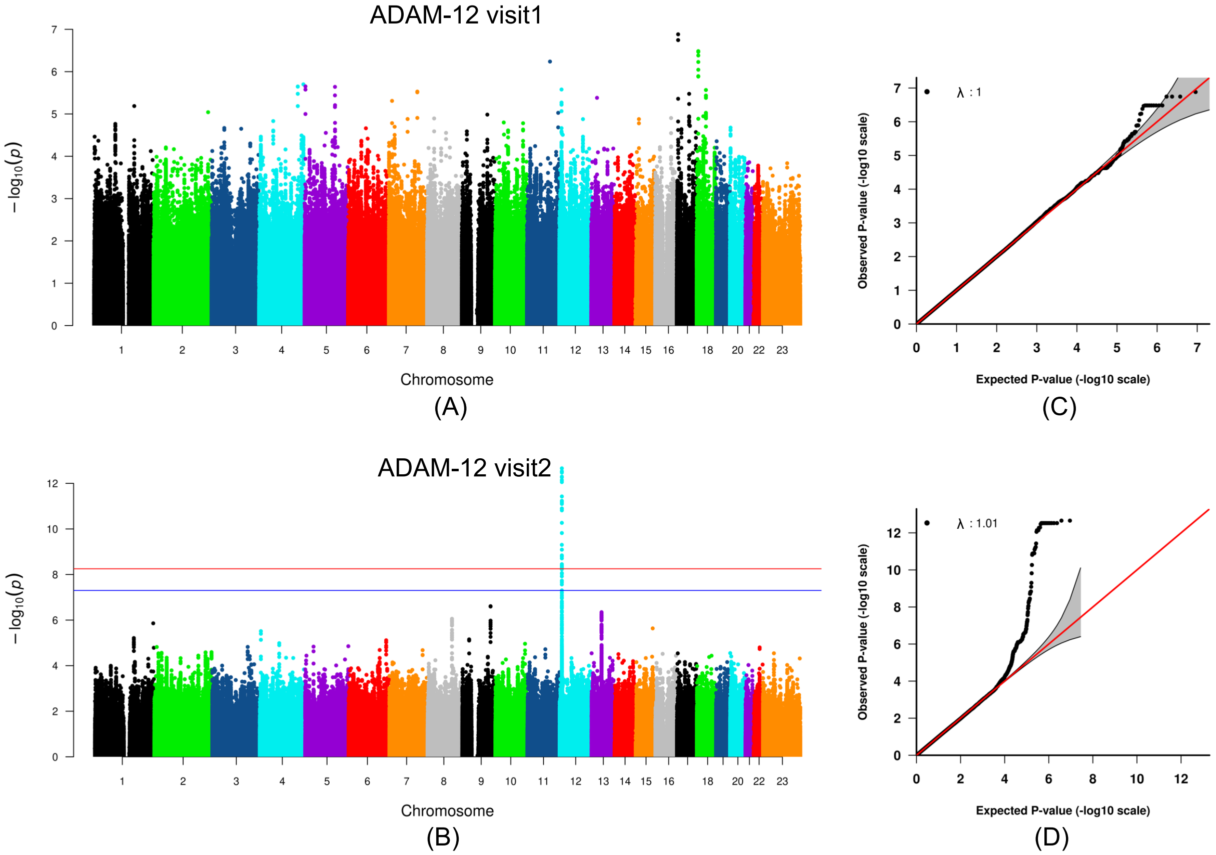

**GWAS of ADAM-12 using individuals of white ancestry.** Manhattan plot displays the associations between SNPs across the genome and a specific trait, with the spikes indicating regions of significant associations. (A) and (B) Manhattan plots for visit1 and visit2 analyses. The chromosomal position of each SNP is displayed along the x-axis and the negative log10 of the association *P*-value is displayed on the y-axis. The red line represents the genome-wide significance level ($P < 5.6\times10^{-9}$) and blue line represents the suggestive significance level ($P < 5\times10^{-8}$). (C) and (D) The quantile-quantile plots for visit1 and visit2 analyses. λ is the genomic control value. The plot displays the distribution of *P*-values against a theoretical distribution, under the null hypothesis of no association. Points (representing SNPs) that align with the diagonal line suggest adherence to the null hypothesis, with a genomic control value near one indicating no inflation. Upward deviations from this line at the higher end indicate SNPs significantly associated with the trait beyond random chance.

**Supplemental Figure 6**

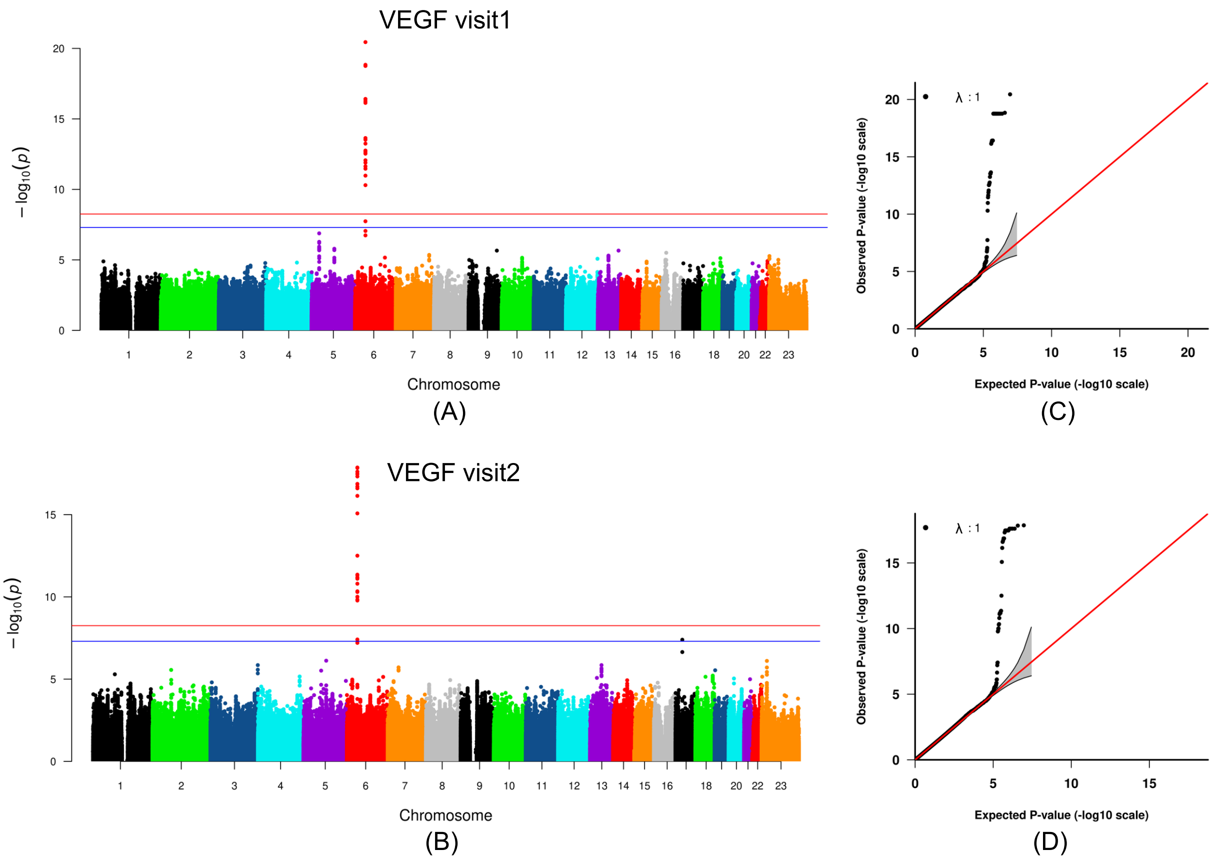

**GWAS of VEGF using individuals of white ancestry.** Manhattan plot displays the associations between SNPs across the genome and a specific trait, with the spikes indicating regions of significant associations. (A) and (B) Manhattan plots for visit1 and visit2 analyses. The chromosomal position of each SNP is displayed along the x-axis and the negative log10 of the association *P*-value is displayed on the y-axis. The red line represents the genome-wide significance level ($P < 5.6\times10^{-9}$) and blue line represents the suggestive significance level ($P < 5\times10^{-8}$). (C) and (D) The quantile-quantile plots for visit1 and visit2 analyses. λ is the genomic control value. The plot displays the distribution of *P*-values against a theoretical distribution, under the null hypothesis of no association. Points (representing SNPs) that align with the diagonal line suggest adherence to the null hypothesis, with a genomic control value near one indicating no inflation. Upward deviations from this line at the higher end indicate SNPs significantly associated with the trait beyond random chance.

**Supplemental Figure 7**

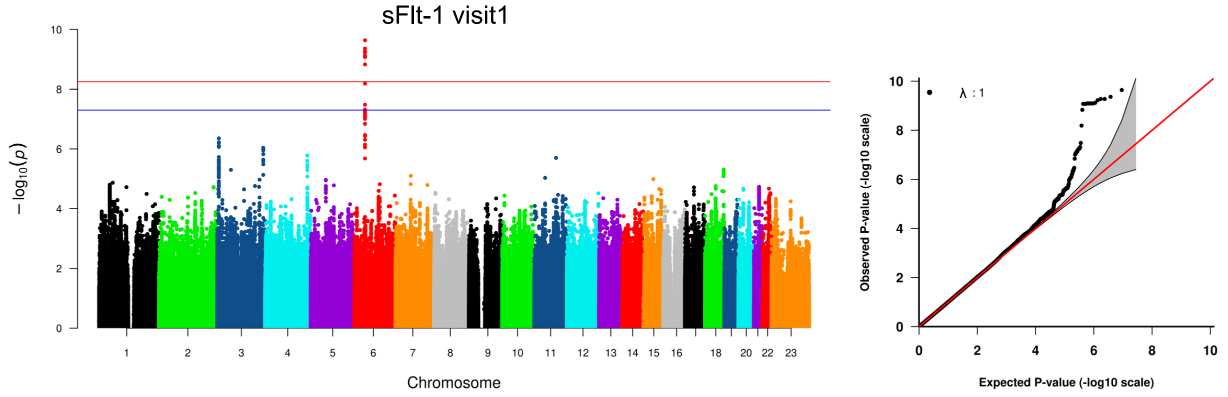

(A) (B)

**GWAS of sFlt-1 using individuals of white ancestry.** Manhattan plot displays the associations between SNPs across the genome and a specific trait, with the spikes indicating regions of significant associations. (A) Manhattan plot for visit1 analysis. The chromosomal position of each SNP is displayed along the x-axis and the negative log10 of the association *P*-value is displayed on the y-axis. The red line represents the genome-wide significance level ($P < 5.6\times10^{-9}$) and blue line represents the suggestive significance level ($P < 5\times10^{-8}$). (B) The quantile-quantile plot for visit1 analysis. λ is the genomic control value. The plot displays the distribution of *P*-values against a theoretical distribution, under the null hypothesis of no association. Points (representing SNPs) that align with the diagonal line suggest adherence to the null hypothesis, with a genomic control value near one indicating no inflation. Upward deviations from this line at the higher end indicate SNPs significantly associated with the trait beyond random chance.

**Supplementary Figure 8**

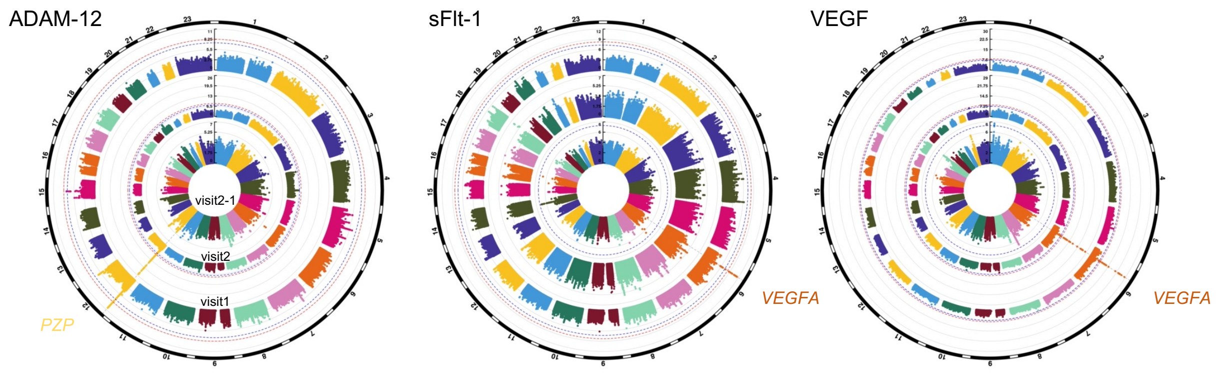

**Circular Manhattan plots of ADAM-12, sFlt-1, and VEGF after additional adjustment for gestational age at the time of blood collection.** Manhattan plot displays the associations between SNPs across the genome and a specific trait, with the spikes indicating regions of significant associations. This circular format presents results from multiple GWAS simultaneously. The chromosomal position of each single SNP is displayed along the circle and the negative log10 of the association *P*-value is displayed on the radius. The red line represents the genome-wide significance level ($P < 5.6\times10^{-9}$) and blue line represents the suggestive significance level ($P < 5\times10^{-8}$). Results for visit1 are displayed on the outer circle, visit2 on the middle circle, and visit2-1 on the inner circle.

**Supplemental Figure 9**

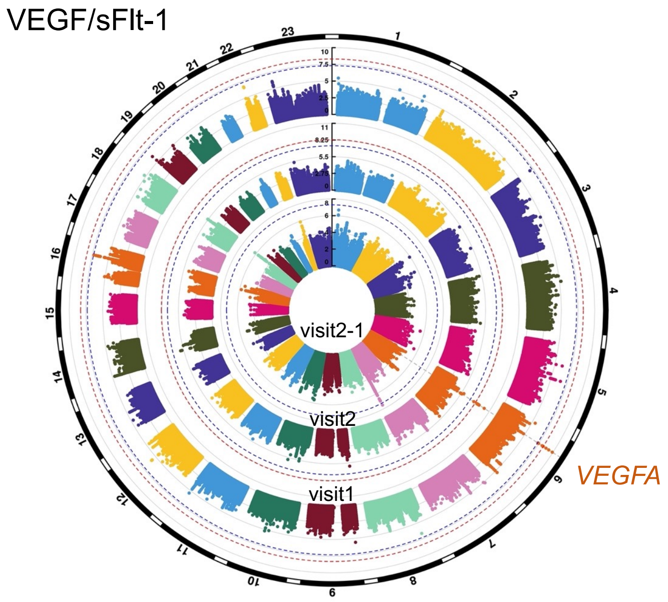

**Circular Manhattan plot of VEGF/sFlt-1.** Manhattan plot displays the associations between SNPs across the genome and a specific trait, with the spikes indicating regions of significant associations. This circular format presents results from multiple GWAS simultaneously. The chromosomal position of each single SNP is displayed along the circle and the negative log10 of the association *P*-value is displayed on the radius. The red line represents the genome-wide significance level ($P < 5\times10^{-9} = 5\times10^{-8}/10$, considering VEGF/sFlt-1 as a new 10$^{th}$ protein) and blue line represents the suggestive significance level ($P < 5\times10^{-8}$). Results for visit1 are displayed on the outer circle, visit2 on the middle circle, and visit2-1 on the inner circle.

**Supplementary Figure 10**

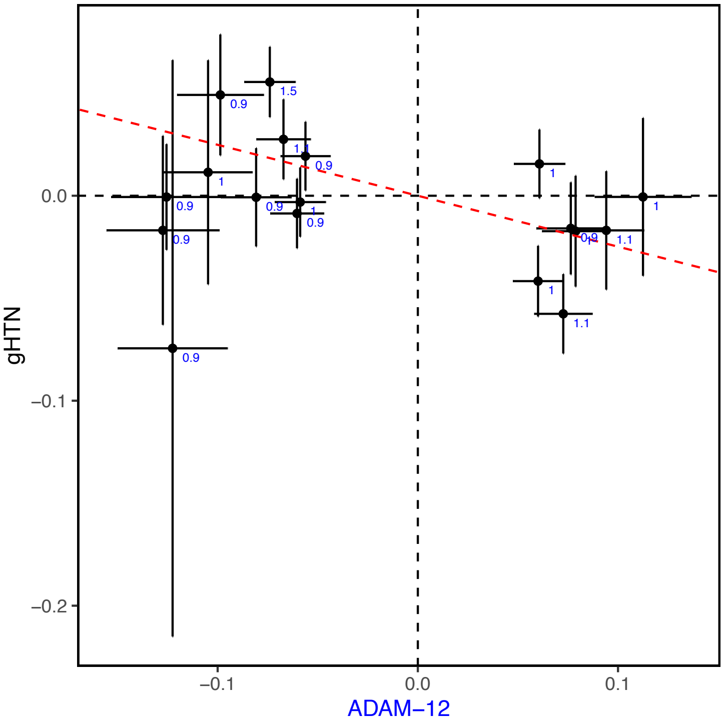

**Scatter plot of genetic effects on gHTN plotted against genetic effects on ADAM-12 levels at visit1.** Each black dot represents a SNP used as an instrumental variable (IV). The selected IV SNPs, which are independent, have $P < 1\times10^{-5}$ in the GWAS of ADAM-12. The whiskers represent standard errors of estimated genetic effects, and the red dashed line shows the estimated effects from MR-RAPS. The blue values are the proportions (%) of ADAM-12 variance explained by individual IV SNPs.

### Supplemental Figure 11

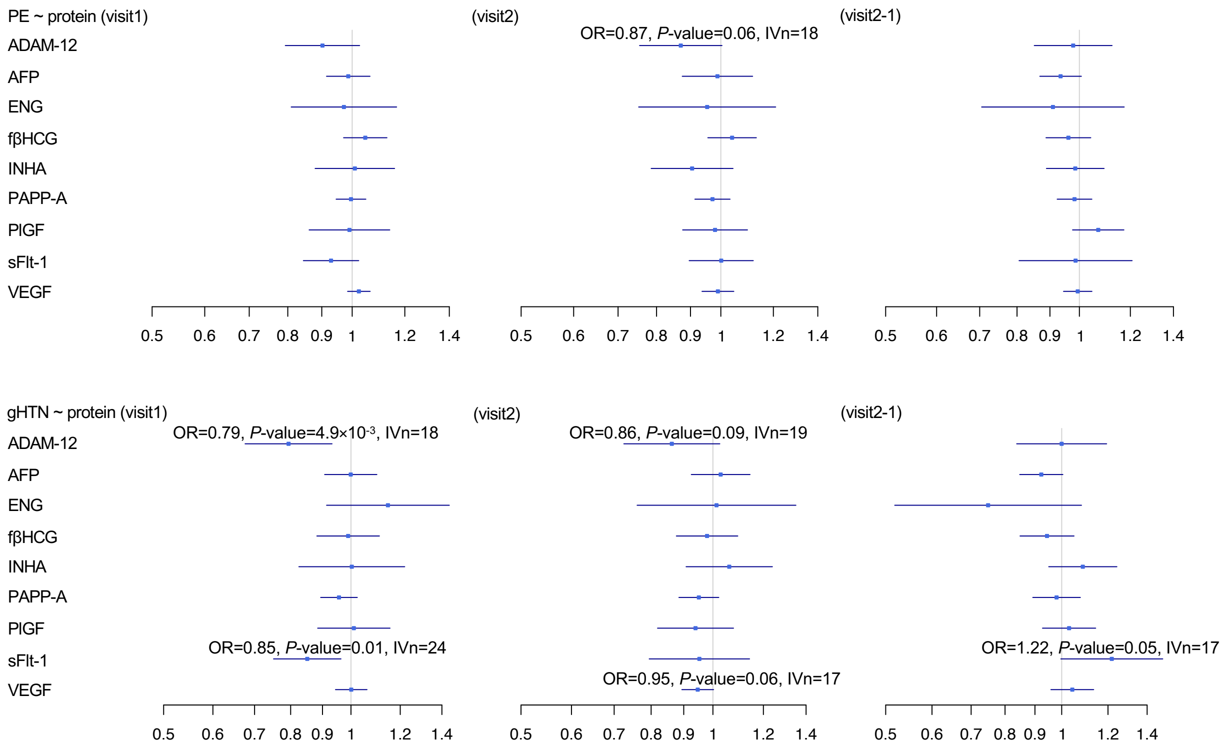

**Causal estimates of the serum levels of nine placental proteins on PE and gHTN.** *P*-values were determined by the two-sample IVW method. The squares represent the causal estimates on the odds ratio (OR) scale, and the whiskers show the corresponding 95% confidence intervals.

### Supplemental Figure 12

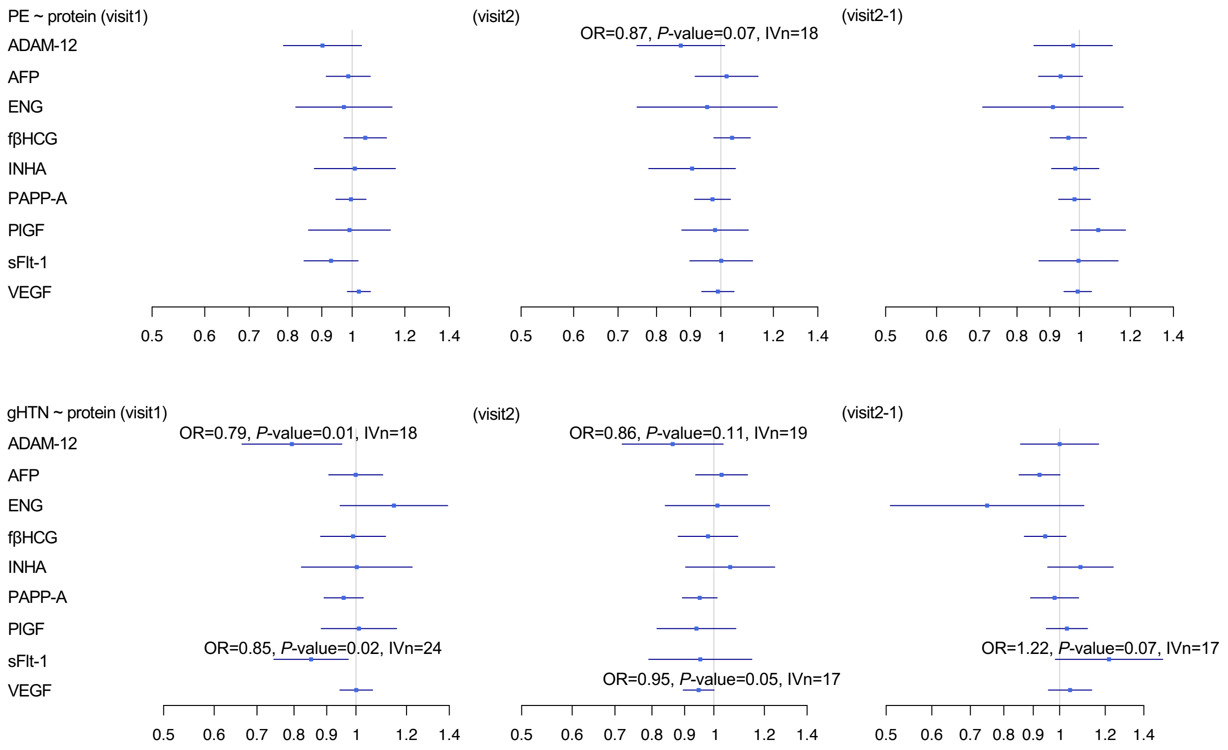

**Causal estimates of the serum levels of nine placental proteins on PE and gHTN.** *P*-values were determined by the two-sample MR-PRESSO method. The squares represent the causal estimates on the odds ratio (OR) scale, and the whiskers show the corresponding 95% confidence intervals.

**Supplementary Figure 13**

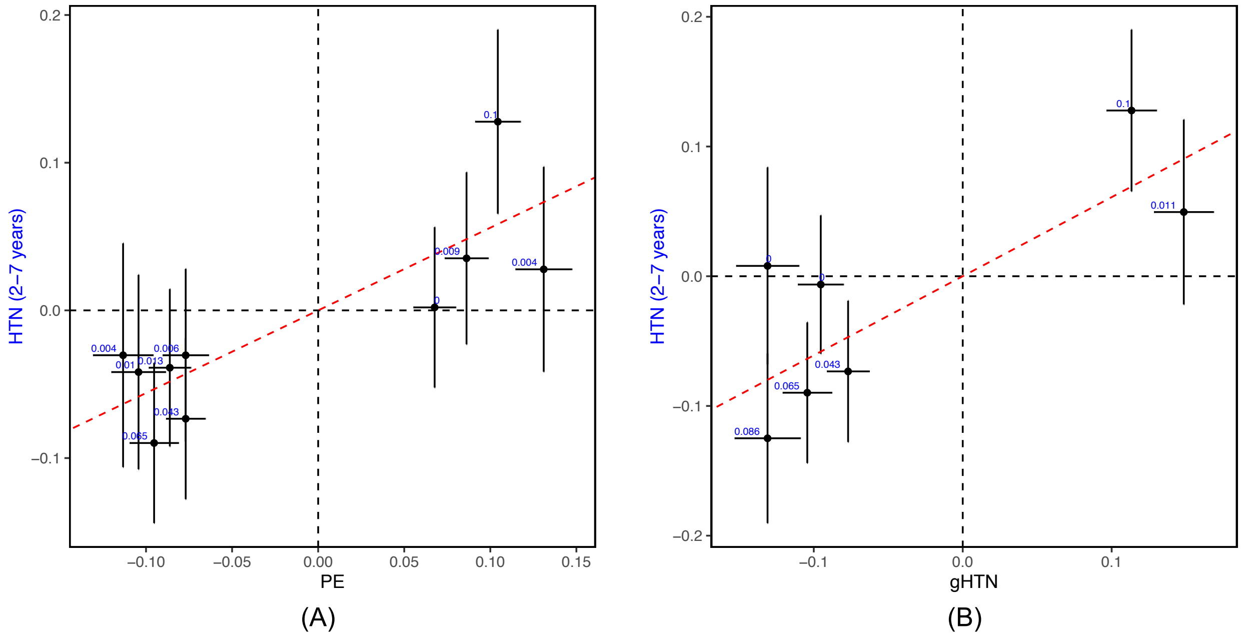

**Scatter plots of genetic effects on long-term postpartum HTN (2-7 years) plotted against genetic effects on PE and gHTN.** (A) PE as the exposure; and (B) gHTN as the exposure. Each black dot represents a SNP used as an instrumental variable (IV). The selected IV SNPs, which are independent, have $P < 5\times10^{-8}$ from a recent multi-ancestry meta-analysis of GWAS of PE/gHTN. The whiskers represent standard errors of estimated genetic effects, and the red dashed line shows the estimated effects from MR-RAPS. The blue values are the proportions (%) of long-term postpartum HTN variance explained by individual IV SNPs.

**Supplemental Figure 14**

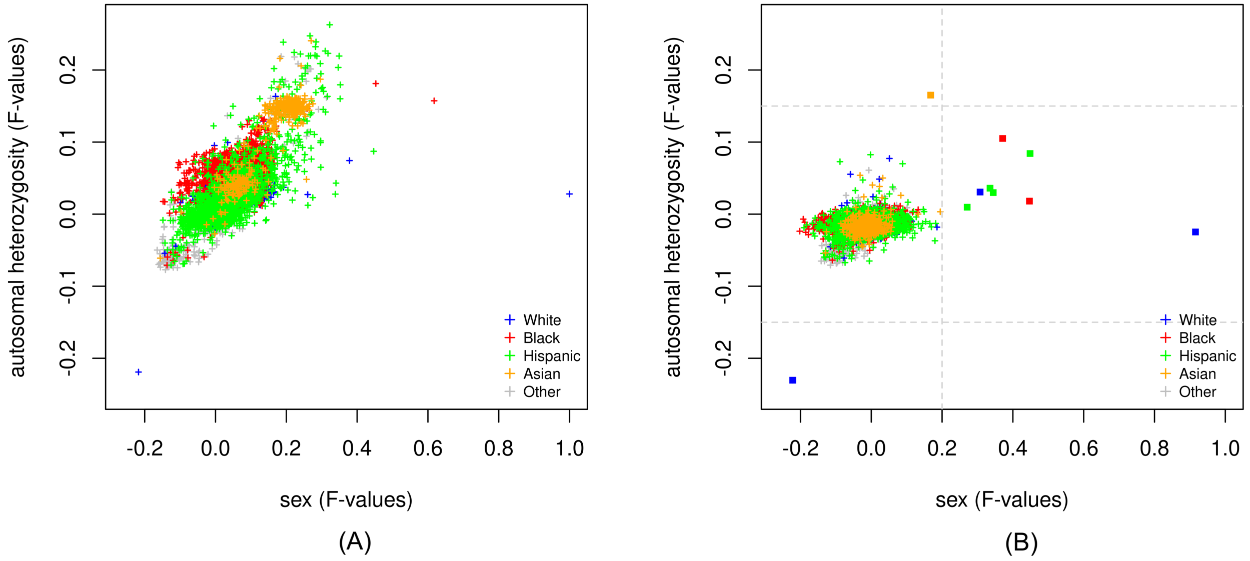

**Assessment of sex concordance and autosomal heterozygosity in the nuMoM2b cohort.** (A) No principal component (PC) adjustment. (B) PC-adjusted results. Each dot represents an individual, and the dots are color-coded according to self-reported race.

**Supplemental Figure 15**

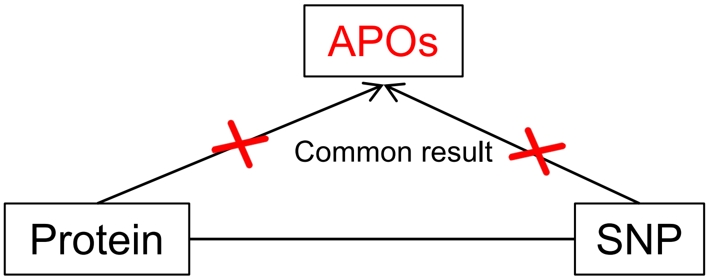

**Illustration of collider bias.** Adjusting for the adverse pregnancy outcome (APO) status in the Protein~SNP regression model may introduce collider bias if both the protein and SNP are independent causes of APO status.

**Supplemental Table 1**

Race of nuMoM2b individuals and nuMoM2b-HHS individuals

| **Baseline Characteristics** | **nuMoM2b (n=10038)** | **nuMoM2b-HHS (n=4484)** |
|---|---|---|
| *Maternal race n (%)* | | |
| White non-Hispanic | 5989 (59.7%) | 2786 (62.1%) |
| Black non-Hispanic | 1418 (14.1%) | 618 (13.8%) |
| Hispanic | 1700 (16.9%) | 735 (16.4%) |
| Asian | 407 (4.1%) | 135 (3.0%) |
| Other | 524 (5.2%) | 210 (4.7%) |

### Supplemental Table 2

Timing of data collection in nuMoM2b and nuMoM2b-HHS[1, 2]

| Question Domains, Samples, and Clinical Evaluations | Pregnancy Trimester (nuMoM2b)* | | | | Postpartum (nuMoM2b-HHS) | |
|---|---|---|---|---|---|---|
| | Visit1 | Visit2 | Visit3 | Delivery | Interval Contact | In-person visit |
| Demographic characteristics | X | X | X | X | X | X |
| Medical history | X | X | X | X | X | X |
| Psychological factors | X | X | X | | | |
| Biometric measurements | X | X | X | | | X |
| Ultrasound | X | X | X | | | |
| Biospecimens | | | | | | |
| Urine | X | X | X | | | X |
| Blood (whole blood, plasma, serum) | X | X | X | X | | X |
| Cervicovaginal fluid | X | X | X | X | | |
| Cord blood (whole blood, plasma, serum), Neonatal saliva | | | | X | | |
| Placenta, fetal membranes, umbilical cord segment | | | | X | | |
| Symptoms or diagnoses between the visit3 and the admission for delivery | | | | X | | |
| Participant assessment of delivery route/reasons | | | | X | | |

* Study visits were during the following gestational age intervals: first trimester, 6 weeks 0 days to 13 weeks 6 days; second trimester, 16 weeks 0 days to 21 weeks 6 days; and third trimester, 22 weeks 0 days to 29 weeks 6 days.

1. HAAS DM, PARKER CB, WING DA, et al. A description of the methods of the Nulliparous Pregnancy Outcomes Study: monitoring mothers-to-be (nuMoM2b). Am J Obstet Gynecol 2015;212:539 e1-39 e24.
2. HAAS DM, EHRENTHAL DB, KOCH MA, et al. Pregnancy as a Window to Future Cardiovascular Health: Design and Implementation of the nuMoM2b Heart Health Study. Am J Epidemiol 2016;183:519-30.

**Supplemental Table 3**

The heritability estimates in percentage (%) from Genome-wide Complex Trait Analysis (GCTA)

| | Visit 1 | Visit 2 | Visit2-1 |
|---|---|---|---|
| ADAM-12 | NA* | 21.7±12.4 | NA |
| AFP | NA | NA | NA |
| ENG | 30.2±12.8 | 16.7±11.8 | NA |
| fβHCG | 14.5±9.2 | NA | NA |
| INHA | 19.6±11.9 | NA | NA |
| PAPP-A | NA | 21.4±9.9 | NA |
| PlGF | 24.6±12.8 | 28±12.7 | NA |
| sFlt-1 | 29±12.4 | 20.7±10.3 | 31±13.9 |
| VEGF | 16.3±11.3 | NA | 19.9±14 |

The values represent the variance explained by genome ± standard error (%)

* NA denotes that the analysis was not properly converged

**Supplemental Table 4**

SNPs achieving suggestive genome-wide significance ($P$-value < $5\times10^{-8}$) in the GWAS of placental protein levels

| SNP | Chr | Position | A1 | A2 | A1_freq | Effect | *P*-value |
|---|---|---|---|---|---|---|---|
| ADAM-12 (visit1) | | | | | | | |
| rs4316551 | 12 | 9326362 | T | G | 0.35 | -0.07 | 6.8E-9 |
| rs7952890 | 12 | 9325722 | G | C | 0.35 | -0.07 | 6.9E-9 |
| rs4316550 | 12 | 9326244 | A | G | 0.35 | -0.07 | 8.8E-9 |
| rs4262771 | 12 | 9326722 | C | G | 0.36 | -0.07 | 8.9E-9 |
| rs4589351 | 12 | 9326275 | T | C | 0.35 | -0.07 | 1.6E-8 |
| rs4370983 | 12 | 9326310 | G | C | 0.23 | -0.08 | 2.7E-8 |
| ADAM-12 (visit2) | | | | | | | |
| rs6487735 | 12 | 9278806 | T | C | 0.47 | -0.10 | 3.0E-22 |
| rs7960104 | 12 | 9283043 | C | T | 0.47 | -0.10 | 6.8E-22 |
| rs10743634 | 12 | 9280392 | T | G | 0.47 | -0.10 | 7.2E-22 |
| rs10743636 | 12 | 9281464 | C | T | 0.47 | -0.10 | 7.6E-22 |
| rs10771464 | 12 | 9280254 | G | A | 0.47 | -0.10 | 1.0E-21 |
| rs1549428 | 12 | 9282256 | A | G | 0.49 | 0.10 | 1.3E-21 |
| rs1549429 | 12 | 9282246 | T | C | 0.49 | 0.10 | 1.3E-21 |
| rs2113899 | 12 | 9280887 | T | C | 0.49 | 0.10 | 1.3E-21 |
| rs7135251 | 12 | 9282219 | T | G | 0.49 | 0.10 | 1.3E-21 |
| rs7300369 | 12 | 9281942 | A | G | 0.47 | -0.10 | 1.5E-21 |
| rs2113900 | 12 | 9280830 | A | T | 0.47 | -0.10 | 3.3E-21 |
| rs10743632 | 12 | 9280307 | A | G | 0.49 | -0.10 | 3.6E-21 |
| rs10771463 | 12 | 9280193 | C | G | 0.47 | -0.10 | 3.7E-21 |
| rs61916194 | 12 | 9283487 | T | C | 0.49 | -0.10 | 5.8E-21 |
| rs61916193 | 12 | 9283485 | T | C | 0.49 | -0.10 | 6.7E-21 |
| rs10743633 | 12 | 9280356 | G | A | 0.49 | -0.10 | 6.9E-21 |
| rs1549426 | 12 | 9282648 | T | C | 0.49 | -0.10 | 7.6E-21 |
| rs6487747 | 12 | 9282978 | C | T | 0.49 | -0.10 | 7.6E-21 |
| rs6487748 | 12 | 9283172 | G | A | 0.49 | -0.10 | 7.6E-21 |
| rs1549427 | 12 | 9282344 | A | G | 0.49 | -0.09 | 1.1E-20 |
| rs1549430 | 12 | 9281009 | G | A | 0.49 | -0.09 | 1.1E-20 |
| rs4514480 | 12 | 9281387 | T | C | 0.49 | -0.09 | 1.1E-20 |
| rs7300172 | 12 | 9281827 | A | G | 0.49 | -0.09 | 1.1E-20 |
| rs397850695 | 12 | 9279174 | A | AT | 0.49 | -0.09 | 7.2E-20 |
| rs12366847 | 12 | 9311372 | A | G | 0.40 | -0.09 | 1.2E-19 |
| rs7960183 | 12 | 9310761 | C | T | 0.40 | -0.09 | 1.2E-19 |
| rs12317441 | 12 | 9310388 | G | A | 0.33 | -0.10 | 9.5E-19 |
| rs7957287 | 12 | 9318173 | G | A | 0.43 | -0.09 | 2.7E-18 |
| rs6487734 | 12 | 9274924 | G | C | 0.37 | -0.09 | 3.8E-18 |
| rs10843408 | 12 | 9319861 | T | C | 0.43 | -0.09 | 5.3E-18 |
| rs10843404 | 12 | 9318619 | C | G | 0.43 | -0.09 | 5.4E-18 |
| rs10843400 | 12 | 9318235 | T | C | 0.42 | -0.09 | 7.0E-18 |
| rs4271436 | 12 | 9320019 | A | G | 0.43 | -0.09 | 7.1E-18 |
| rs4271437 | 12 | 9320047 | A | G | 0.41 | -0.09 | 7.7E-18 |
| rs11050218 | 12 | 9320023 | A | G | 0.43 | -0.09 | 8.3E-18 |
| rs10743661 | 12 | 9319428 | T | C | 0.41 | -0.09 | 1.1E-17 |
| rs10743660 | 12 | 9319419 | T | C | 0.41 | -0.09 | 1.2E-17 |
| rs34337 | 12 | 9271712 | G | A | 0.38 | -0.09 | 1.9E-17 |
| rs1059171 | 12 | 9323351 | G | A | 0.42 | -0.09 | 8.7E-17 |
| rs1059172 | 12 | 9323348 | T | C | 0.42 | -0.09 | 8.7E-17 |
| rs7952890 | 12 | 9325722 | G | C | 0.35 | -0.08 | 7.1E-14 |
| rs4370983 | 12 | 9326310 | G | C | 0.23 | -0.09 | 9.5E-14 |
| rs4316551 | 12 | 9326362 | T | G | 0.35 | -0.08 | 1.5E-13 |
| rs4316550 | 12 | 9326244 | A | G | 0.35 | -0.08 | 1.6E-13 |
| rs12369816 | 12 | 9254686 | C | T | 0.31 | -0.09 | 1.7E-13 |
| rs10843422 | 12 | 9324656 | T | G | 0.32 | -0.08 | 1.9E-13 |
| rs11049845 | 12 | 9260846 | T | C | 0.31 | -0.08 | 2.7E-13 |

| | | | | | | | |
|---|---|---|---|---|---|---|---|
| rs11049846 | 12 | 9261011 | G | A | 0.31 | -0.08 | 2.7E-13 |
| rs4262771 | 12 | 9326722 | C | G | 0.36 | -0.08 | 3.8E-13 |
| rs4589351 | 12 | 9326275 | T | C | 0.35 | -0.08 | 5.0E-13 |
| rs7298028 | 12 | 9303444 | C | T | 0.49 | 0.07 | 6.4E-13 |
| rs10743654 | 12 | 9305929 | A | G | 0.50 | -0.07 | 6.6E-13 |
| rs2911825 | 12 | 9307302 | T | G | 0.50 | -0.07 | 1.2E-12 |
| rs9971685 | 12 | 9307258 | C | G | 0.50 | -0.07 | 1.2E-12 |
| rs11049781 | 12 | 9247292 | T | C | 0.31 | -0.08 | 1.3E-12 |
| rs10492110 | 12 | 9340896 | A | G | 0.29 | -0.08 | 2.0E-12 |
| rs12298908 | 12 | 9173646 | T | A | 0.23 | 0.08 | 7.7E-12 |
| rs10771539 | 12 | 9326861 | C | G | 0.43 | 0.07 | 8.4E-12 |
| rs2377762 | 12 | 9313503 | T | C | 0.50 | -0.07 | 8.9E-12 |
| rs12321232 | 12 | 9163072 | T | G | 0.22 | 0.08 | 1.9E-11 |
| rs4322447 | 12 | 9326791 | T | G | 0.27 | -0.08 | 2.0E-11 |
| rs201046098 | 12 | 9304044 | A | C | 0.38 | 0.07 | 3.1E-11 |
| rs10843050 | 12 | 9178009 | C | T | 0.23 | 0.08 | 3.3E-11 |
| rs2277413 | 12 | 9165188 | G | A | 0.30 | 0.07 | 3.3E-11 |
| rs397973429 | 12 | 9324515 | C | CT | 0.47 | 0.07 | 4.7E-11 |
| rs2059759 | 12 | 9244342 | G | A | 0.31 | -0.08 | 4.7E-11 |
| rs252024 | 12 | 9269572 | A | G | 0.39 | -0.07 | 5.6E-11 |
| rs7299515 | 12 | 9322198 | G | A | 0.47 | 0.07 | 8.5E-11 |
| rs7311982 | 12 | 9162261 | T | C | 0.32 | 0.07 | 8.8E-11 |
| rs6487821 | 12 | 9321124 | T | C | 0.46 | 0.07 | 9.9E-11 |
| rs10771532 | 12 | 9321808 | C | T | 0.47 | 0.07 | 1.1E-10 |
| rs61917373 | 12 | 9302746 | G | A | 0.38 | 0.07 | 1.4E-10 |
| rs7954451 | 12 | 9243525 | T | G | 0.23 | -0.08 | 1.7E-10 |
| rs4883237 | 12 | 9324123 | A | G | 0.50 | -0.07 | 1.8E-10 |
| rs6487824 | 12 | 9321286 | G | T | 0.47 | 0.07 | 1.9E-10 |
| rs35276849 | 12 | 9244269 | C | CT | 0.29 | -0.07 | 2.0E-10 |
| rs1035848 | 12 | 9209508 | G | T | 0.26 | -0.08 | 2.1E-10 |
| rs10771531 | 12 | 9321709 | C | A | 0.48 | 0.07 | 2.1E-10 |
| rs7954383 | 12 | 9321608 | T | C | 0.48 | 0.07 | 2.1E-10 |
| rs4883238 | 12 | 9324245 | A | G | 0.47 | 0.07 | 2.6E-10 |
| rs10843222 | 12 | 9241470 | A | T | 0.22 | -0.08 | 3.2E-10 |
| rs7311758 | 12 | 9321129 | A | G | 0.47 | 0.06 | 3.3E-10 |
| rs11049626 | 12 | 9225517 | G | A | 0.26 | -0.07 | 4.4E-10 |
| rs12303039 | 12 | 9237702 | A | G | 0.25 | -0.07 | 7.5E-10 |
| rs55809356 | 12 | 9311178 | T | G | 0.36 | 0.07 | 8.6E-10 |
| rs10843223 | 12 | 9241903 | T | C | 0.22 | -0.08 | 8.9E-10 |
| rs7971371 | 12 | 9326586 | G | A | 0.36 | 0.07 | 1.5E-9 |
| rs3741848 | 12 | 9242426 | C | T | 0.27 | -0.07 | 1.9E-9 |
| rs4353323 | 12 | 9211628 | G | A | 0.27 | -0.07 | 2.2E-9 |
| rs10843160 | 12 | 9215694 | A | G | 0.27 | -0.07 | 2.5E-9 |
| rs7137569 | 12 | 9219539 | T | C | 0.27 | -0.07 | 3.5E-9 |
| rs71656520 | 12 | 9331369 | C | CTGGAGCAGG | 0.45 | 0.06 | 3.5E-9 |
| rs2377747 | 12 | 9217768 | T | C | 0.27 | -0.07 | 4.0E-9 |
| rs7137281 | 12 | 9227282 | A | G | 0.27 | -0.07 | 4.8E-9 |
| rs76611603 | 12 | 9228049 | C | CACTT | 0.27 | -0.07 | 4.8E-9 |
| rs12580730 | 12 | 9310189 | G | C | 0.39 | 0.06 | 5.5E-9 |
| rs2195208 | 12 | 9207884 | A | G | 0.27 | -0.07 | 5.7E-9 |
| rs2003610 | 12 | 9337211 | T | A | 0.35 | -0.06 | 6.4E-9 |
| rs11612935 | 12 | 9355559 | C | T | 0.33 | 0.06 | 6.5E-9 |
| rs7959473 | 12 | 9333129 | A | G | 0.46 | 0.06 | 7.4E-9 |
| rs4636721 | 12 | 9334981 | T | C | 0.36 | -0.06 | 8.6E-9 |
| rs7958717 | 12 | 9332510 | A | G | 0.46 | 0.06 | 8.8E-9 |
| rs11050312 | 12 | 9339311 | G | T | 0.35 | -0.06 | 8.8E-9 |
| rs7974095 | 12 | 9332511 | T | A | 0.46 | 0.06 | 9.9E-9 |
| rs3741847 | 12 | 9242430 | T | C | 0.26 | -0.07 | 1.1E-8 |
| rs2003859 | 12 | 9337154 | C | A | 0.36 | -0.06 | 1.1E-8 |
| rs34331 | 12 | 9256128 | T | C | 0.18 | 0.08 | 1.5E-8 |
| rs4141479 | 12 | 9335707 | C | T | 0.36 | -0.06 | 2.0E-8 |
| rs397775325 | 12 | 9336794 | TA | T | 0.36 | -0.06 | 2.2E-8 |

| | | | | | | | |
|---|---|---|---|---|---|---|---|
| rs12366431 | 12 | 9364768 | A | G | 0.33 | -0.06 | 2.3E-8 |
| rs7972572 | 12 | 9336366 | A | C | 0.37 | -0.06 | 2.5E-8 |
| rs4883241 | 12 | 9340686 | T | C | 0.35 | -0.06 | 2.7E-8 |
| rs11050283 | 12 | 9336026 | A | G | 0.37 | -0.06 | 2.9E-8 |
| rs9788250 | 12 | 9336207 | G | A | 0.37 | -0.06 | 2.9E-8 |
| rs12828464 | 12 | 9325142 | A | G | 0.38 | 0.06 | 3.7E-8 |
| rs34723854 | 12 | 9203265 | TA | T | 0.27 | -0.06 | 4.3E-8 |
| VEGF (visit1) | | | | | | | |
| rs6921438 | 6 | 43957870 | A | G | 0.45 | -0.36 | 7.9E-30 |
| rs12205248 | 6 | 43958482 | C | T | 0.42 | -0.35 | 2.2E-27 |
| rs13206436 | 6 | 43958041 | A | G | 0.42 | -0.35 | 2.2E-27 |
| rs4349808 | 6 | 43957037 | C | T | 0.42 | -0.35 | 2.2E-27 |
| rs4513773 | 6 | 43957789 | G | A | 0.42 | -0.35 | 2.2E-27 |
| rs4637627 | 6 | 43957590 | A | G | 0.42 | -0.35 | 2.2E-27 |
| rs7763440 | 6 | 43958971 | A | G | 0.42 | -0.35 | 2.2E-27 |
| rs9472168 | 6 | 43961248 | G | A | 0.41 | -0.34 | 5.2E-26 |
| rs11757888 | 6 | 43964582 | T | C | 0.42 | -0.34 | 1.3E-25 |
| rs11757868 | 6 | 43964496 | T | C | 0.42 | -0.34 | 1.4E-25 |
| rs9472172 | 6 | 43963248 | T | C | 0.42 | -0.34 | 1.6E-25 |
| rs11757903 | 6 | 43964486 | A | G | 0.42 | -0.33 | 4.9E-25 |
| rs4320361 | 6 | 43960774 | T | G | 0.46 | -0.33 | 6.3E-25 |
| rs4349809 | 6 | 43957093 | G | T | 0.46 | -0.33 | 7.9E-25 |
| rs4413611 | 6 | 43957026 | A | G | 0.46 | -0.33 | 7.9E-25 |
| rs7767396 | 6 | 43959313 | G | A | 0.46 | -0.33 | 7.9E-25 |
| rs13206012 | 6 | 43965026 | A | G | 0.36 | -0.32 | 5.9E-22 |
| rs9472159 | 6 | 43951958 | A | C | 0.39 | -0.27 | 1.7E-16 |
| rs9472170 | 6 | 43961684 | G | C | 0.39 | 0.25 | 1.6E-14 |
| rs4714719 | 6 | 43964875 | C | T | 0.47 | 0.24 | 2.2E-14 |
| rs4481426 | 6 | 43960371 | C | T | 0.39 | 0.25 | 4.6E-14 |
| rs7745184 | 6 | 43958901 | T | G | 0.41 | 0.24 | 9.9E-14 |
| rs4382251 | 6 | 43957636 | T | C | 0.39 | 0.24 | 2.9E-13 |
| rs9369434 | 6 | 43950670 | T | C | 0.34 | -0.25 | 4.9E-13 |
| rs7745183 | 6 | 43958898 | T | G | 0.43 | 0.22 | 1.4E-12 |
| rs9472158 | 6 | 43951160 | G | A | 0.49 | 0.23 | 2.2E-12 |
| rs9654590 | 6 | 43969052 | C | T | 0.18 | 0.29 | 2.9E-12 |
| rs9472171 | 6 | 43963225 | A | G | 0.41 | 0.22 | 1.0E-11 |
| rs943075 | 6 | 43954468 | A | G | 0.36 | 0.22 | 6.4E-11 |
| rs6916314 | 6 | 43951425 | G | A | 0.48 | 0.21 | 1.0E-10 |
| rs73422214 | 6 | 43954866 | G | A | 0.21 | 0.26 | 6.0E-10 |
| rs9462949 | 6 | 43963610 | G | A | 0.35 | 0.21 | 2.7E-9 |
| rs6916540 | 6 | 43951679 | C | T | 0.49 | 0.19 | 4.1E-9 |
| rs7739450 | 6 | 43943861 | A | G | 0.41 | -0.19 | 7.6E-9 |
| rs7017991 | 8 | 18549017 | G | C | 0.05 | 0.41 | 2.7E-8 |
| rs58397113 | 8 | 18540457 | C | G | 0.05 | 0.40 | 4.4E-8 |
| VEGF (visit2) | | | | | | | |
| rs6921438 | 6 | 43957870 | A | G | 0.47 | -0.32 | 2.5E-28 |
| rs4349808 | 6 | 43957037 | C | T | 0.44 | -0.31 | 2.1E-26 |
| rs4513773 | 6 | 43957789 | G | A | 0.44 | -0.31 | 2.5E-26 |
| rs12205248 | 6 | 43958482 | C | T | 0.44 | -0.31 | 2.6E-26 |
| rs13206436 | 6 | 43958041 | A | G | 0.44 | -0.31 | 2.6E-26 |
| rs7763440 | 6 | 43958971 | A | G | 0.44 | -0.31 | 2.6E-26 |
| rs4637627 | 6 | 43957590 | A | G | 0.44 | -0.31 | 2.8E-26 |
| rs9472168 | 6 | 43961248 | G | A | 0.43 | -0.30 | 1.9E-25 |
| rs4349809 | 6 | 43957093 | G | T | 0.47 | -0.30 | 2.3E-25 |
| rs4413611 | 6 | 43957026 | A | G | 0.47 | -0.30 | 2.3E-25 |
| rs7767396 | 6 | 43959313 | G | A | 0.47 | -0.30 | 3.0E-25 |
| rs4320361 | 6 | 43960774 | T | G | 0.47 | -0.30 | 4.1E-25 |
| rs11757903 | 6 | 43964486 | A | G | 0.44 | -0.30 | 8.7E-25 |
| rs11757888 | 6 | 43964582 | T | C | 0.44 | -0.30 | 1.0E-24 |
| rs9472172 | 6 | 43963248 | T | C | 0.44 | -0.30 | 1.2E-24 |
| rs11757868 | 6 | 43964496 | T | C | 0.44 | -0.30 | 1.6E-24 |
| rs13206012 | 6 | 43965026 | A | G | 0.37 | -0.29 | 4.0E-22 |

| | | | | | | | |
|---|---|---|---|---|---|---|---|
| rs9472159 | 6 | 43951958 | A | C | 0.41 | -0.25 | 8.9E-18 |
| rs9369434 | 6 | 43950670 | T | C | 0.36 | -0.24 | 1.0E-15 |
| rs9472158 | 6 | 43951160 | G | A | 0.47 | 0.23 | 5.8E-15 |
| rs4481426 | 6 | 43960371 | C | T | 0.38 | 0.21 | 1.1E-12 |
| rs7739450 | 6 | 43943861 | A | G | 0.42 | -0.21 | 1.2E-12 |
| rs4382251 | 6 | 43957636 | T | C | 0.38 | 0.21 | 1.3E-12 |
| rs4714719 | 6 | 43964875 | C | T | 0.47 | 0.20 | 1.4E-12 |
| rs9472170 | 6 | 43961684 | G | C | 0.38 | 0.21 | 1.6E-12 |
| rs943075 | 6 | 43954468 | A | G | 0.36 | 0.22 | 1.9E-12 |
| rs7745184 | 6 | 43958901 | T | G | 0.40 | 0.20 | 9.3E-12 |
| rs6916314 | 6 | 43951425 | G | A | 0.46 | 0.20 | 1.0E-11 |
| rs7745183 | 6 | 43958898 | T | G | 0.42 | 0.19 | 4.7E-11 |
| rs6916540 | 6 | 43951679 | C | T | 0.48 | 0.19 | 5.8E-11 |
| rs9462949 | 6 | 43963610 | G | A | 0.35 | 0.20 | 1.1E-10 |
| rs9472171 | 6 | 43963225 | A | G | 0.41 | 0.19 | 1.5E-10 |
| rs9381268 | 6 | 43966158 | T | C | 0.44 | 0.17 | 1.2E-8 |
| rs5951549 | X | 22349075 | C | T | 0.07 | 0.32 | 1.9E-8 |
| rs729391 | 6 | 43950155 | C | T | 0.32 | 0.17 | 4.0E-8 |
| rs9472167 | 6 | 43960924 | A | G | 0.06 | 0.34 | 4.9E-8 |
| sFlt-1 (visit1) | | | | | | | |
| rs4349809 | 6 | 43957093 | G | T | 0.49 | -0.09 | 2.9E-12 |
| rs4413611 | 6 | 43957026 | A | G | 0.49 | -0.09 | 2.9E-12 |
| rs4637627 | 6 | 43957590 | A | G | 0.46 | -0.09 | 3.0E-12 |
| rs4513773 | 6 | 43957789 | G | A | 0.46 | -0.09 | 3.7E-12 |
| rs4349808 | 6 | 43957037 | C | T | 0.46 | -0.09 | 3.7E-12 |
| rs7767396 | 6 | 43959313 | G | A | 0.49 | -0.09 | 3.9E-12 |
| rs4320361 | 6 | 43960774 | T | G | 0.49 | -0.09 | 4.4E-12 |
| rs12205248 | 6 | 43958482 | C | T | 0.46 | -0.09 | 5.0E-12 |
| rs13206436 | 6 | 43958041 | A | G | 0.46 | -0.09 | 5.0E-12 |
| rs7763440 | 6 | 43958971 | A | G | 0.46 | -0.09 | 5.0E-12 |
| rs6921438 | 6 | 43957870 | A | G | 0.48 | -0.09 | 9.9E-12 |
| rs9369434 | 6 | 43950670 | T | C | 0.36 | -0.09 | 1.1E-11 |
| rs11757868 | 6 | 43964496 | T | C | 0.45 | -0.09 | 1.2E-11 |
| rs11757888 | 6 | 43964582 | T | C | 0.45 | -0.09 | 1.8E-11 |
| rs9472172 | 6 | 43963248 | T | C | 0.45 | -0.09 | 2.2E-11 |
| rs9472168 | 6 | 43961248 | G | A | 0.44 | -0.09 | 2.5E-11 |
| rs11757903 | 6 | 43964486 | A | G | 0.45 | -0.09 | 2.9E-11 |
| rs4382251 | 6 | 43957636 | T | C | 0.37 | 0.09 | 1.7E-10 |
| rs4481426 | 6 | 43960371 | C | T | 0.37 | 0.08 | 3.3E-10 |
| rs9472170 | 6 | 43961684 | G | C | 0.37 | 0.08 | 3.4E-10 |
| rs7745183 | 6 | 43958898 | T | G | 0.41 | 0.08 | 7.4E-10 |
| rs13206012 | 6 | 43965026 | A | G | 0.39 | -0.08 | 1.8E-9 |
| rs9472159 | 6 | 43951958 | A | C | 0.42 | -0.08 | 1.9E-9 |
| rs7745184 | 6 | 43958901 | T | G | 0.39 | 0.08 | 1.9E-9 |
| rs9472171 | 6 | 43963225 | A | G | 0.39 | 0.08 | 2.6E-9 |
| rs4714719 | 6 | 43964875 | C | T | 0.45 | 0.07 | 3.3E-8 |
| rs6916540 | 6 | 43951679 | C | T | 0.47 | 0.07 | 4.0E-8 |
| VEGF (visit2-1) | | | | | | | |
| rs72886119 | 1 | 24594227 | C | G | 0.43 | 0.16 | 4.2E-8 |
| fβHCG (visit1) | | | | | | | |
| rs981087 | 6 | 87099684 | C | T | 0.48 | -0.10 | 1.1E-8 |
| rs981086 | 6 | 87100023 | T | A | 0.49 | -0.10 | 1.2E-8 |
| PAPP-A (visit1) | | | | | | | |
| rs10458657 | 10 | 74021178 | C | A | 0.39 | 0.19 | 2.9E-8 |

### Supplemental Table 5

Comparison of the top GWAS SNPs using all individuals versus using only individuals of white ancestry

| SNP | Chr | Position | A1 | White | | | | Multi-ethnic | | | |
|---|---|---|---|---|---|---|---|---|---|---|---|
| | | | | n | MAF | Effect | *P*-value | n | MAF | Effect | *P*-value |
| ADAM-12 (visit1) | | | | | | | | | | | |
| rs6487735 | 12 | 9278806 | T | 1307 | 0.49 | -0.06 | $9.5\times10^{-5}$ | 2259 | 0.47 | -0.06 | $2.6\times10^{-7}$ |
| rs2277413 | 12 | 9165188 | G | 1307 | 0.29 | 0.06 | $1.9\times10^{-4}$ | 2259 | 0.3 | 0.06 | $1.1\times10^{-5}$ |
| rs4316551 | 12 | 9326362 | T | 1307 | 0.33 | -0.07 | $5.9\times10^{-6}$ | 2259 | 0.35 | -0.07 | $6.8\times10^{-9}$ |
| VEGF (visit1) | | | | | | | | | | | |
| rs6921438 | 6 | 43957870 | A | 1053 | 0.45 | -0.35 | $3.6\times10^{-21}$ | 1831 | 0.45 | -0.36 | $7.9\times10^{-30}$ |
| rs4349809 | 6 | 43957093 | G | 1053 | 0.44 | -0.34 | $1.7\times10^{-19}$ | 1831 | 0.46 | -0.33 | $7.9\times10^{-25}$ |
| sFlt-1 (visit1) | | | | | | | | | | | |
| rs6921438 | 6 | 43957870 | A | 1308 | 0.48 | -0.1 | $4.3\times10^{-10}$ | 2262 | 0.48 | -0.09 | $9.9\times10^{-12}$ |
| rs4349809 | 6 | 43957093 | G | 1308 | 0.47 | -0.1 | $8.4\times10^{-10}$ | 2262 | 0.49 | -0.09 | $2.9\times10^{-12}$ |
| ADAM-12 (visit2) | | | | | | | | | | | |
| rs6487735 | 12 | 9278806 | T | 1241 | 0.49 | -0.1 | $5.1\times10^{-13}$ | 2085 | 0.47 | -0.1 | $3\times10^{-22}$ |
| rs2277413 | 12 | 9165188 | G | 1241 | 0.3 | 0.09 | $5.9\times10^{-9}$ | 2085 | 0.3 | 0.07 | $3.3\times10^{-11}$ |
| rs4316551 | 12 | 9326362 | T | 1241 | 0.33 | -0.08 | $5\times10^{-9}$ | 2085 | 0.35 | -0.08 | $1.5\times10^{-13}$ |
| VEGF (visit2) | | | | | | | | | | | |
| rs6921438 | 6 | 43957870 | A | 1135 | 0.47 | -0.31 | $1.4\times10^{-18}$ | 1942 | 0.47 | -0.32 | $2.5\times10^{-28}$ |
| rs4349809 | 6 | 43957093 | G | 1135 | 0.45 | -0.31 | $2.4\times10^{-18}$ | 1942 | 0.47 | -0.3 | $2.3\times10^{-25}$ |

**Supplemental Table 6**

Association of SNPs previously published for circulating VEGF levels in the nuMoM2b cohort

| | Chr | Position | A1 | MAF* | Effect* | *P*-value* | Ref |
|---|---|---|---|---|---|---|---|
| *VEGFA* | | | | | | | |
| rs6921438 | 6 | 43957870 | A | 0.45/0.47/0.47 | -/-/- | $7.94\times10^{-30}/2.49\times10^{-28}/2.09\times10^{-171}$ | 1 |
| rs6921438 | 6 | 43957870 | A | 0.45/0.47/0.46 | -/-/- | $7.94\times10^{-30}/2.49\times10^{-28}/7.4\times10^{-1467}$ | 2 |
| rs6921438 | 6 | 43957870 | A | 0.45/0.47/0.49 | -/-/- | $7.94\times10^{-30}/2.49\times10^{-28}/6.11\times10^{-506}$ | 3 |
| rs7767396 | 6 | 43959313 | G | 0.46/0.47/0.48 | -/-/- | $7.91\times10^{-25}/3.01\times10^{-25}/8.35\times10^{-105}$ | 4 |
| *VLDLR* | | | | | | | |
| rs7030781 | 9 | 2686273 | T | 0.48/0.48/0.42 | -/-/- | $0.41/0.49/2.57\times10^{-15}$ | 1 |
| rs2375981 | 9 | 2692583 | C | 0.49/0.49/0.54 | +/+/+ | $0.21/0.25/1.5\times10^{-100}$ | 2 |
| rs10738760 | 9 | 2681186 | A | 0.48/0.48/0.49 | +/+/+ | $0.3/0.46/1.96\times10^{-34}$ | 3 |
| rs7030781 | 9 | 2686273 | T | 0.48/0.48/0.37 | -/-/- | $0.41/0.49/1.57\times10^{-13}$ | 4 |

* The results of visit1 are displayed first, followed by the results of visit2, and finally, the referenced study is presented.

1. AHOLA-OLLI AV, WURTZ P, HAVULINNA AS, et al. Genome-wide Association Study Identifies 27 Loci Influencing Concentrations of Circulating Cytokines and Growth Factors. Am J Hum Genet 2017;100:40-50.
2. CHOI SH, RUGGIERO D, SORICE R, et al. Six Novel Loci Associated with Circulating VEGF Levels Identified by a Meta-analysis of Genome-Wide Association Studies. PLoS Genet 2016;12:e1005874.
3. DEBETTE S, VISVIKIS-SIEST S, CHEN MH, et al. Identification of cis- and trans-acting genetic variants explaining up to half the variation in circulating vascular endothelial growth factor levels. Circ Res 2011;109:554-63.
4. SLIZ E, KALAOJA M, AHOLA-OLLI A, et al. Genome-wide association study identifies seven novel loci associating with circulating cytokines and cell adhesion molecules in Finns. J Med Genet 2019;56:607-16.
